# Outcome and life expectancy associated with novel subgroups of atrial fibrillation: a data-driven cluster analysis

**DOI:** 10.1101/2023.10.07.23296702

**Authors:** Yuefeng Yu, Jiang Li, Ying Sun, Bowei Yu, Xiao Tan, Bin Wang, Yingli Lu, Ningjian Wang

## Abstract

**Aim:** This study aimed to develop a refined classification of atrial fibrillation (AF) according to comprehensive risk factors to identify patients at high-risk for poor prognosis and their life expectancy.

**Method:** A total of 7,391 participants aged 40-69 with AF at baseline, from UK Biobank, were classified into five clusters, based on seven clustering variables including age and six risk factor categories (metabolic disease, respiratory disease, cardiovascular disease, renal/immune-mediated disease, mental health disease, acute illness). Difference in the risk of death and major complications, as well as reductions in life expectancy, among clusters were estimated. Replication was done in 2,399 participants with newly diagnosed AF within two years after baseline.

**Results:** Five distinct AF clusters were identified: acute illness-related, mental health-related, cardiovascular disease-related, immune-and-renal disease-related, and respiratory-and- metabolic disease-related AF. Patients with respiratory-and-metabolic disease-related AF had the highest risk of death, acute myocardial infarction, heart failure, and cerebral ischemic stroke, while those with acute illness-related AF had the lowest corresponding risk. In addition, compared with individuals with acute illness-related AF, those with respiratory-and- metabolic disease-related and mental health-related AF had the top 2 greatest loss of life expectancy. Furthermore, genetic variants for AF had different effect among the five clusters. Replication analysis confirmed the result stability.

**Conclusion:** A novel AF classification was developed, which provided insights into varying life expectancy and risks of death and complications among AF subgroups with distinct characteristics. It offers a practical approach for identifying high-risk patients, which might help to tailor precise interventions for AF management.

## Introduction

Atrial fibrillation (AF), as the most common sustained arrhythmia, imposes a huge burden on individuals and public health worldwide ^1^. In the 2019 global statistics, there were approximately 59.7 million people with AF and atrial flutter and the number is increasing ^2^. Even under existing treatment strategies, patients with AF still had higher risk of mortality, heart failure, and stroke ^3^. One possible explanation for these shortcomings is that the current diagnosis of AF is mainly based on measurement of characteristics of AF episodes including presentation, duration, and spontaneous termination ^4^, but AF is heterogeneous with regard to clinical presentation, progression, and outcomes.

According to the characteristics of AF episodes, the classification subgroups of AF could be first diagnosed as paroxysmal, persistent, and permanent. However, recommendations for AF management do not rely on the temporal AF patterns, except for the restoration of sinus rhythm ^5,6^. Considering that the pathophysiological mechanism and risk factors for AF are complex, multifactorial, and interacting ^7,8^, personalized assessment of the pathophysiological process involved in the individual patient by using clinical characteristics, blood biomarkers, and non-invasive substrate determination may improve classification. Further, the guidelines suggested that personalizing management decisions for patients should be based on a structured characterization of AF, including four AF domains-Stroke risk, Symptom severity, Severity of AF burden, and Substrate severity^9^. Assessment tools have been established for the first three domains (CHA_2_DS_2_-VASC score ^10^ for Stroke risk, EHRA symptom score ^11^ for Symptom severity, ICD code for Severity of AF burden). However, there are no clear quantitative criteria for evaluating Substrate severity.

Our previous study provided comprehensive information about the associations of risk factor profiles with incident AF ^12^. Based on these risk factors, we aimed to set up a refined classification based on unsupervised and data-driven cluster analyses in UK population that could identify subgroups at different risks of different complications, to help enable personalized intervention. (**Supplementary Figure S1**).

## Methods

### Study population

A total of 499,980 adults aged 40-69 years old were recruited from the UK Biobank at baseline^13^. We declare that all data are publicly available in the UK Biobank (UKB) repository^13^. Among them, 8,293 participants were diagnosed with AF before baseline. After excluding 902 participants for missing data on risk factors, 7,391 participants with AF were included in the main analysis (**Supplementary Figure S1**), with a follow-up of 86,860 person-years. Further, 2,399 participants with newly diagnosed AF within two years after baseline were extracted as a cohort for replication analyses.

### Ascertainment of incident AF, AF complications and death

Incident AF cases (both primary and secondary diagnoses) were identified from the “first occurrence of health outcomes defined by International Statistical Classification of Diseases and Related Health Problems, Tenth Revision (ICD-10) (field identification [ID] 131350, ICD-10 I48). Similarly, information on acute myocardial infarction (AMI), heart failure (HF), and cerebral ischemic stroke (CIS) was obtained by corresponding ICD-10 codes (**Supplementary Table S1**). Information on all-cause and cardiovascular deaths of patients was obtained from the death register records.

### Definition of risk factors

The *2020 European Society of Cardiology (ESC) Guidelines for the diagnosis and management of AF developed in collaboration with the European Association for Cardio-Thoracic Surgery* summarized 46 risk factors for incident AF, primarily covering demographic factors, health behaviors, clinical comorbidities, cardiometabolic factors and genetic factors^3^.

After an exclusion for unavailable/inadequate data and information integration, we included 25 risk factors in the present study (**Supplementary Table S2**), covering demographic factors (age, sex, race, education, Townsend deprivation index (TDI), air pollution), health behaviors (current smoking, excessive alcohol intake, physical inactivity, excessive physical activity, loneliness, broad depression), clinical comorbidities (hypertension, diabetes/prediabetes, overweight/obesity, low low-density lipoprotein cholesterol (LDL-C), low triglycerides, elevated CRP, sleep apnea, obstructive pulmonary disease (COPD), renal dysfunction, cardiovascular conditions, immune mediated disease, acute illness) and genetic factor (genetic risk score). Detailed information on rationale for risk factor selection and definition of individual risk factors in UK Biobank dataset has been described in our previous study^12^.

### Selection of risk factors

We used the Least Absolute Shrinkage and Selection Operator (LASSO) regression with 10-fold cross-validated^14^ to screen for the principal risk factors for AF, among all 25 factors. To facilitate easy interpretation, continuous variables were coded as binary categorical variables based on clinical significance. Details were shown in **Supplementary Method 1**. We chose the largest lambda as possible to obtain the leanest model while ensuring a good-fitting model **(Supplementary Figures S2 and S3)**. 23 factors with nonzero coefficient were selected for subsequent analyses (**Supplementary Figure S1 and Table S3**).

### Categorization of risk factors

Because of the complexity of AF onset and the interaction of risk factors, we used factor analysis to categorize risk factors ^15^. Given that four non-modifiable factors (age, sex race, GRS) had a relatively independent role in AF occurrence, and three social-level factors (education, TDI, air pollution) were difficult to change on patients’ own, we excluded these seven risk factors in factor analysis. Then, we grouped the remaining 16 risk factors into 6 categories (**Supplementary Figure 1, Supplementary Method 2**). Category 1 was named as metabolic factors, including hypertension, overweight/obesity, and diabetes/prediabetes. Category 2 was named as respiratory factors, including COPD, sleep apnea, current smoking, and elevated CRP. Category 3 was named as cardiovascular factors, including cardiovascular diseases, low levels of LDL-C and triglycerides. Category 4 was named as renal and immunity diseases, including renal dysfunction, immune-mediated disease, and excessive alcohol intake. Category 5 was named as mental health, including loneliness and broad depression. Category 6 was named as acute illness. Finally, we established a score of each category for all participants by summing all risk factors within it, to replace the values obtained in the factor analysis considering the convenience of clinical application (**Supplementary Table S4**). A higher score indicated a higher risk of the corresponding risk profile.

### Cluster analysis

To focus on risk factors at the individual levels, the 3 social-level risk factors mentioned above were also not to be considered as cluster variables. Sex and race were excluded, as binary variables are not fit for cluster analysis. GRS is not a routinely available variable and was excluded to increase the generalizability of the algorithm. Finally, age and six category scores were chosen for cluster variables, and sex, race, GRS, and 3 social risk factors were included as covariates in the following analyses (**Supplementary Figure S1**). Cluster variables were standardized to a mean of 0 and standard deviation (SD) of 1 before cluster analysis. Consensus clustering analysis was employed to estimate the optimal cluster number^16^. We specified a range of cluster numbers (K=2-12), and iteratively applied the consensus clustering algorithm for each K value. K of 5 was determined as the optimal for the clustering model. More details were shown in **Supplementary Method 3**. The cluster stability was assessed by Jaccard means, which reflected the consistency of participants in a cluster among 1000 times random resampling.

### Characterization and prognostic analysis

Cohen’s effect size (d) was calculated to indicate the standardized difference of characters between each two clusters. The risk of death and complications was calculated using Cox regression, adjusting sex and race in the basic model, and further adjusting GRS, education, TDI, and air pollution in the additional model. Years of life lost were calculated as the difference in the areas under the survival curves of each cluster ^17–19^. Hazard ratios for death at specified age (interactions between age and cluster were added in Cox regression), prevalence of each AF cluster by age, and population all-cause mortality rates (driven from the Office for National Statistics ^20^) were used to estimate reductions in life expectancy associated with AF cluster^17^. In addition, we also used 491,291 participants without AF within two years after baseline as the reference in the supplementary analysis.

In addition, we examined the risk of death and complications, and years of life lost for each AF cluster, compared with participants without AF in 2 years follow-up. Associations between clusters and genotypes were calculated with PLINK2.0 ^21^, after adjustment of age, sex, assessment centers, top 10 principal components of ancestry, and dominance-deviation. To increase the practicability of the clustering algorithm and validate its replicability in newly-onset AF cohorts. First, we established clustering centers for the five clusters in the main analysis. Second, to verify the accuracy of these clustering centers, we re-clustered the participants in the main analysis according to the shortest Pearson distance to the above clustering center. Third, we further classified newly-onset AF patients who developed AF within 2 years of follow-up using established clustering centers. Finally, we assessed the characteristics of these five clusters and the differences in the risk of death and complications among them. Detailed algorithm of clustering AF patients was available on GitHub (https://github.com/JiangLi-1/clustercnp).

## Results

### Distribution of features by clusters

We employed the K-means algorithm to perform clustering on participants with AF at baseline. The Jaccard similarity coefficients in all clusters consistently exceeded 0.76. Demographic, anthropometric, and clinical data for the five clusters are shown in **Table 1**. The five clusters exhibited distinctive patterns, which were characterized by the standardized means of the cluster variables (**Figure 1**). Cluster 1, including 587 (7.9%) participants, was characterized by young age and most likely to have acute illnesses, and labelled as acute illness-related AF. Cluster 2, including 1,593 (21.6%), was characterized by poor mental health and moderately likely to have acute illnesses, and labelled as mental health-related AF. Cluster 3 was labelled as cardiovascular disease (CVD)-related AF, including 2,912 (39.4%) participants and characterized by high prevalence of CVD and acute illnesses. Cluster 4 was labelled as immune-and-renal disease related AF, including 1,253 (17.0%) participants and characterized by high prevalence of immune disease and renal dysfunction. The 1,046 (14.2%) participants in cluster 5 (labelled as respiratory-and-metabolic disease-related AF) were characterized by prominent respiratory and metabolic problems. The pairwise comparisons of the clustering variables between clusters were shown in **Supplementary Figure S4**. Most of the results achieved Bonferroni corrected statistical significance (*P* < 0.005).

**Table 1.**
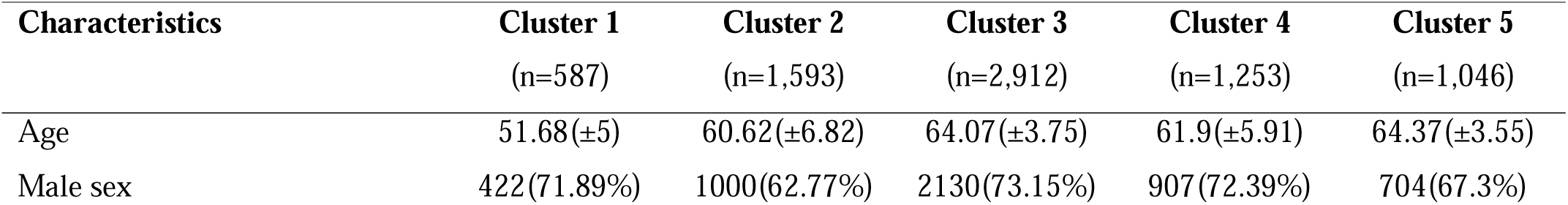
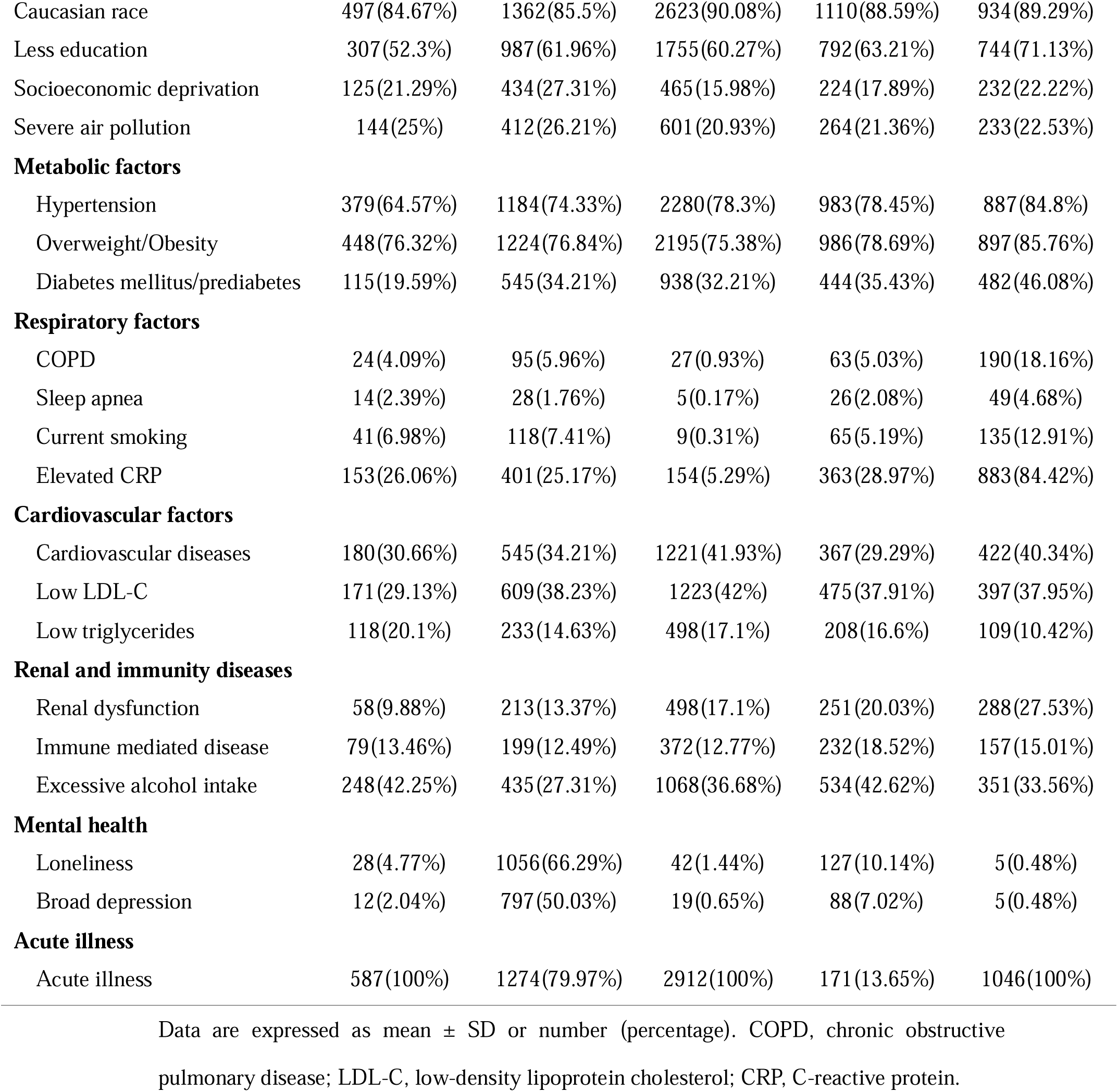
Basic characteristics for the five clusters.

**Figure 1.**
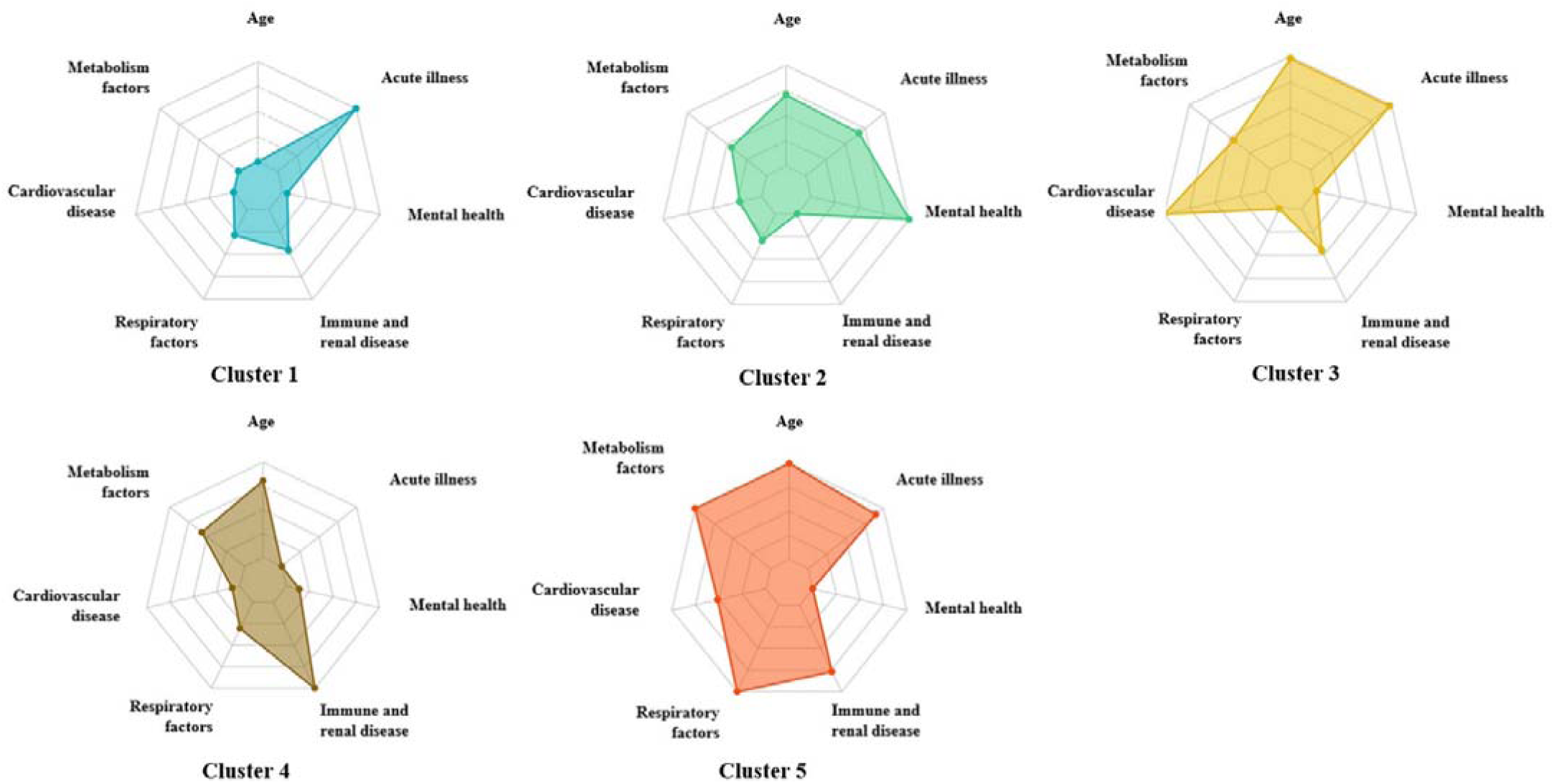
Cluster features differed among five clusters. All the values of cluster features were centered to a mean value of 0 and an SD of 1. All the negative values were converted to positive values by adding a fixed value to ensure that no cluster feature was below 0.

### Risk of death and major complications among AF clusters

During mean follow-up of 11.8 (SD 2.8) years, 1462 (19.8%) deaths were recorded. The risk of all-cause mortality gradually increased from cluster 1 to cluster 5. Compared with participants in cluster 1, the risk of death was increased stepwise for those in other clusters (cluster 2 to 5: HR, 95%CI 1.85, 1.40-2.43; 1.95, 1.49-2.53; 1.95, 1.47-2.58; 3.21, 2.44-4.23), after adjusting sex and race (**Figure 2 and Supplementary Table S5-S6**). For cardiovascular death, we found the similar trend as all-cause death (**Figure 2 and Supplementary Table S5-S6**).

**Figure 2.**
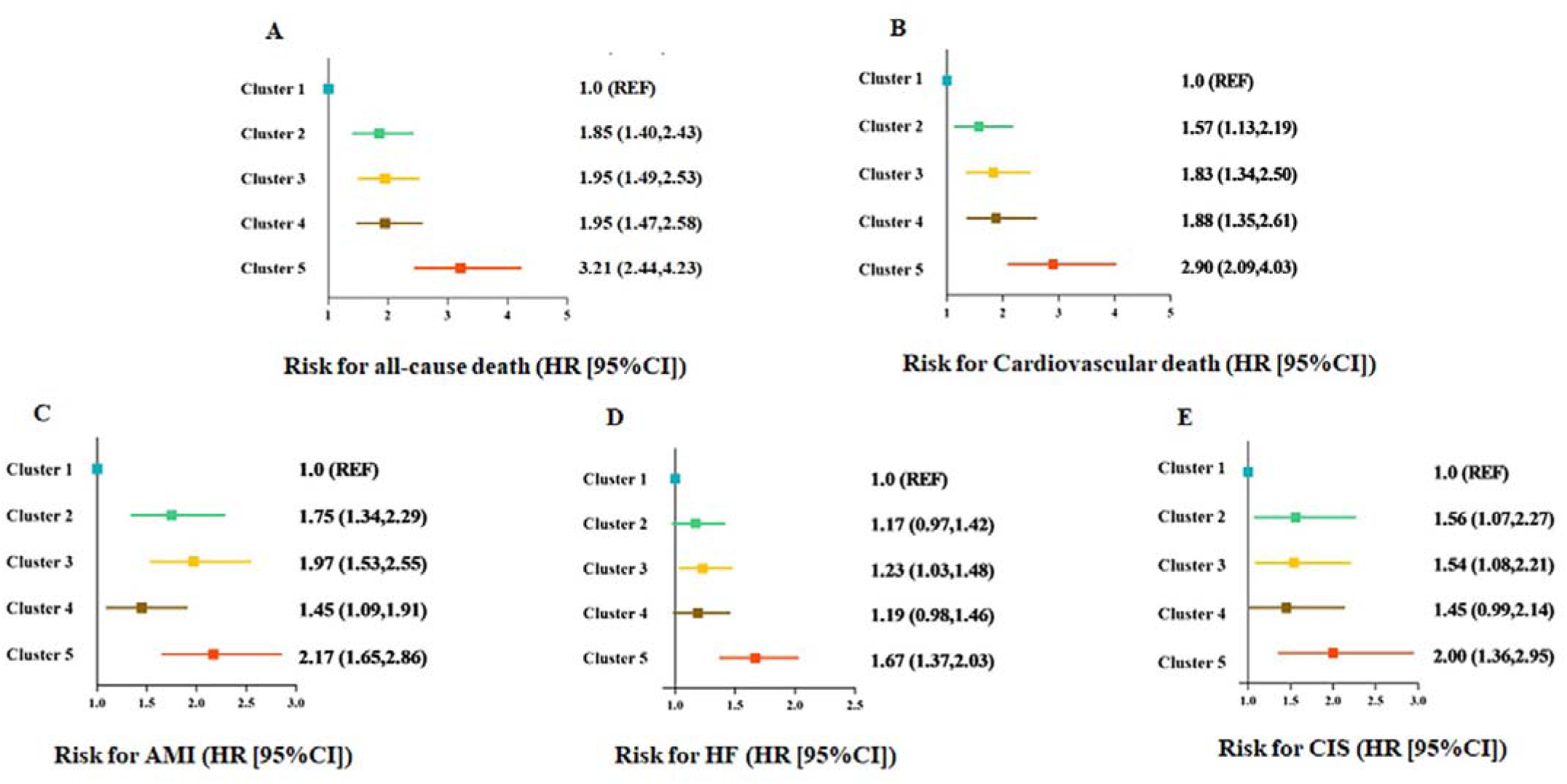
Difference in risk of death and major complications among five clusters. Hazard ratios of all-cause death, cardiovascular death, AMI, HF and CIS for AF cluster 2 to 4, comparing with cluster 1. Cox proportional hazards models were adjusted for sex and race. AMI, acute myocardial infarction, HF, heart failure, CIS, cerebral ischemic stroke.

The association between AF clusters and major complications was shown in Figure 2 and Supplementary Table S7. Compared with participants in cluster 1, those in other clusters had significantly higher risk of AMI, stepwise increased from cluster 4 (1.45, 1.09-1.91), cluster 2 (1.75, 1.34-2.29), cluster 3 (1.97, 1.53-2.55), to cluster 5 (2.17, 1.65-2.86). Compared with participants in cluster 1, those in cluster 3 and 5 were at significantly higher risk of HF (cluster 3 and 5: 1.23, 1.03-1.48; 1.67, 1.37-2.03). Additionally, cluster 2, 3 and 5 were at significantly higher risk of CIS (cluster 2, 3 and 5: 1.56, 1.07-2.27; 1.54, 1.08-2.21; 2.00, 1.36-2.95), compared with cluster 1. After further adjusting for GRS, education, TDI, and air pollution, the results were similar (**Supplementary Table S8**).

### Life expectancy among AF clusters

Compared with participants in cluster 1, reduction of life expectancy stepwise increased from cluster 4, cluster 3, cluster 2 and cluster 5 **(Figure 3, Supplementary Table S9**). For example, at the age of 40 years, we projected a life loss of 32.6 years (95%CI 31.1-34.1) and 30.3 (29.0-31.9) for participants in cluster 5 and cluster 2, respectively, which was significantly higher than those in other two clusters.

**Figure 3.**
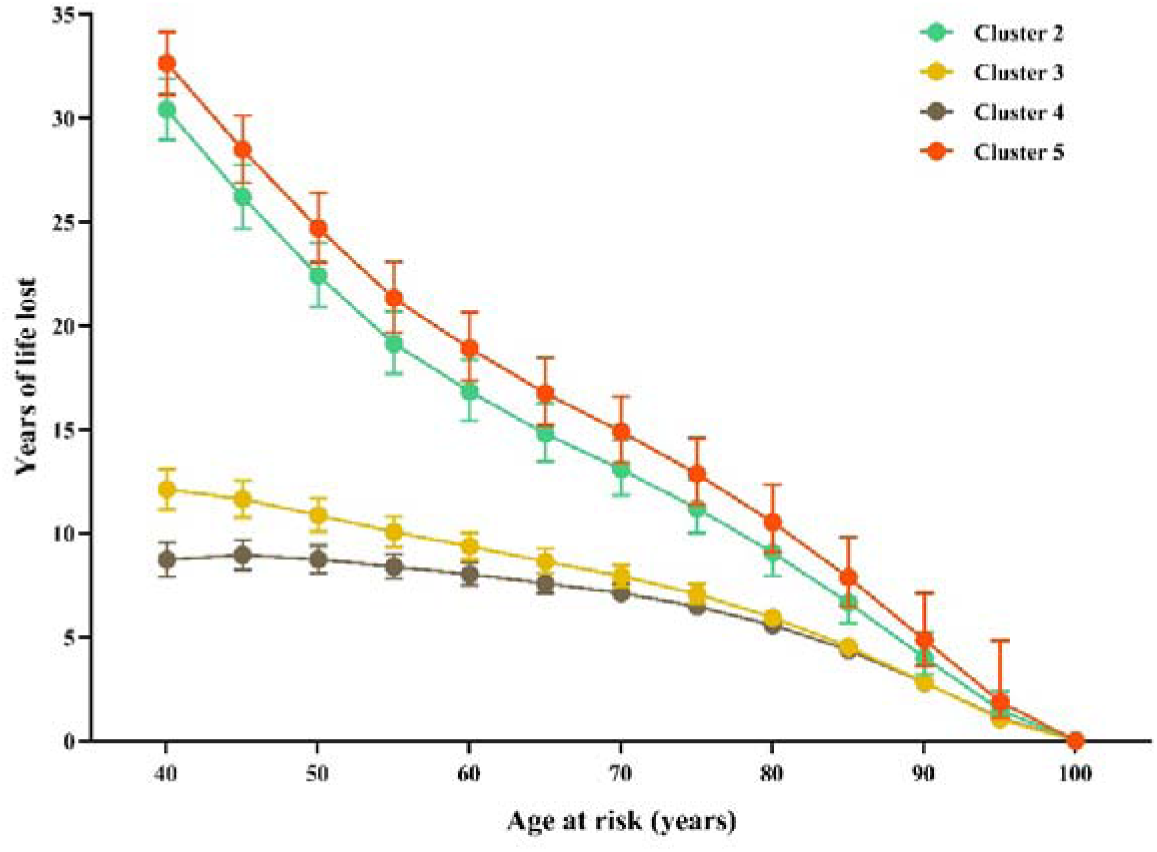
Reductions of life expectancy for participants in AF cluster 2 to 4, comparing with cluster 1. The model was adjusted for sex and race. Cluster 2; yellow, cluster 3; brown, cluster 4; red, cluster 5.

### Cluster migration patterns from clinical characteristics to new clusters

The Sankey diagram shows the patterns of redistribution from classification of clinical characteristics to new clusters (**Figure 4**). Types of AF under the ICD classification had different tendencies in new clusters (**Supplementary Table S10)**. Paroxysmal AF preferred to be classified in cluster 3 (CVD-related AF) and 4 (immune-and-renal disease related AF) (36.1% and 22.3%). However, persistent and chronic AF were prone to be classified into cluster 2 (mental health-related AF) and 3 (immune-and-renal disease related AF) (persistent AF: 39.7% and 17.2%; chronic AF: 31.3% and 25%). Notably, unspecified AF by clinic characters were redistributed into different clusters.

**Figure 4.**
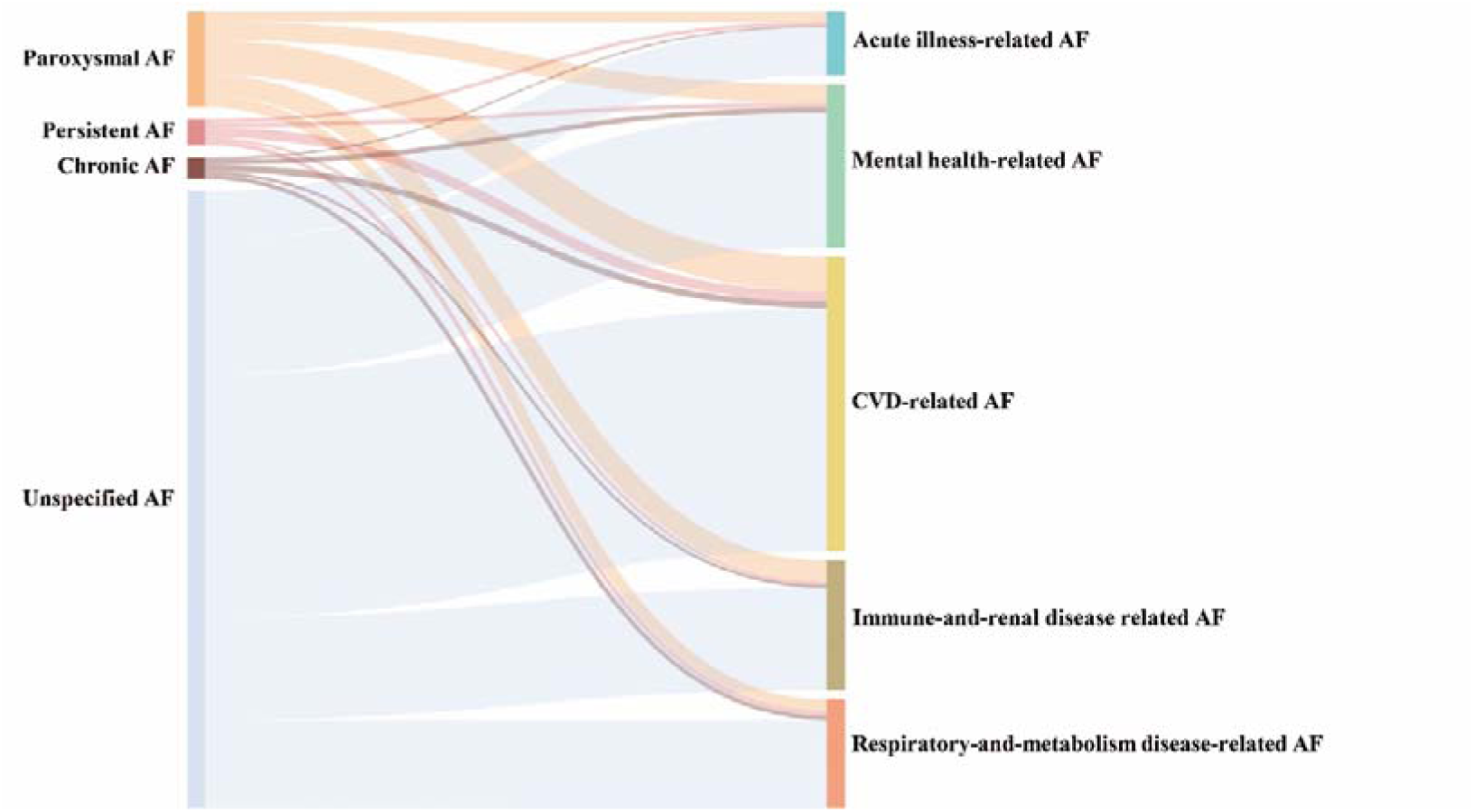
Cluster migration pattern from clinical diagnosis to five clusters

### Associations of outcome, life expectancy using non-AF participants as the reference

Compared with non-AF participants, all clusters had significantly higher risk of death and major complications (**Supplementary Table S11)**. Among three complications, the risk of HF was highest for all clusters. Compared with non-AF participants, individuals in cluster 2 and 5 at age 40 years were projected to die about 30 years earlier than those without AF in 2 years follow-up, and the numbers in cluster 1, 3 and 4 were about 10 years (**Figure 5 and Supplementary Table S12**). In addition, we analysed the association of genetic loci risk for AF ^22^ with each AF cluster, compared with non-AF participants (**Supplementary Table S13)**. The association of three genetic variants (rs11264280, rs2129977, rs2359171) with at least one cluster reached the genome-wide level of significance (P < 5*10^-8) and differed among five clusters (**Table 2**).

**Table 2.**
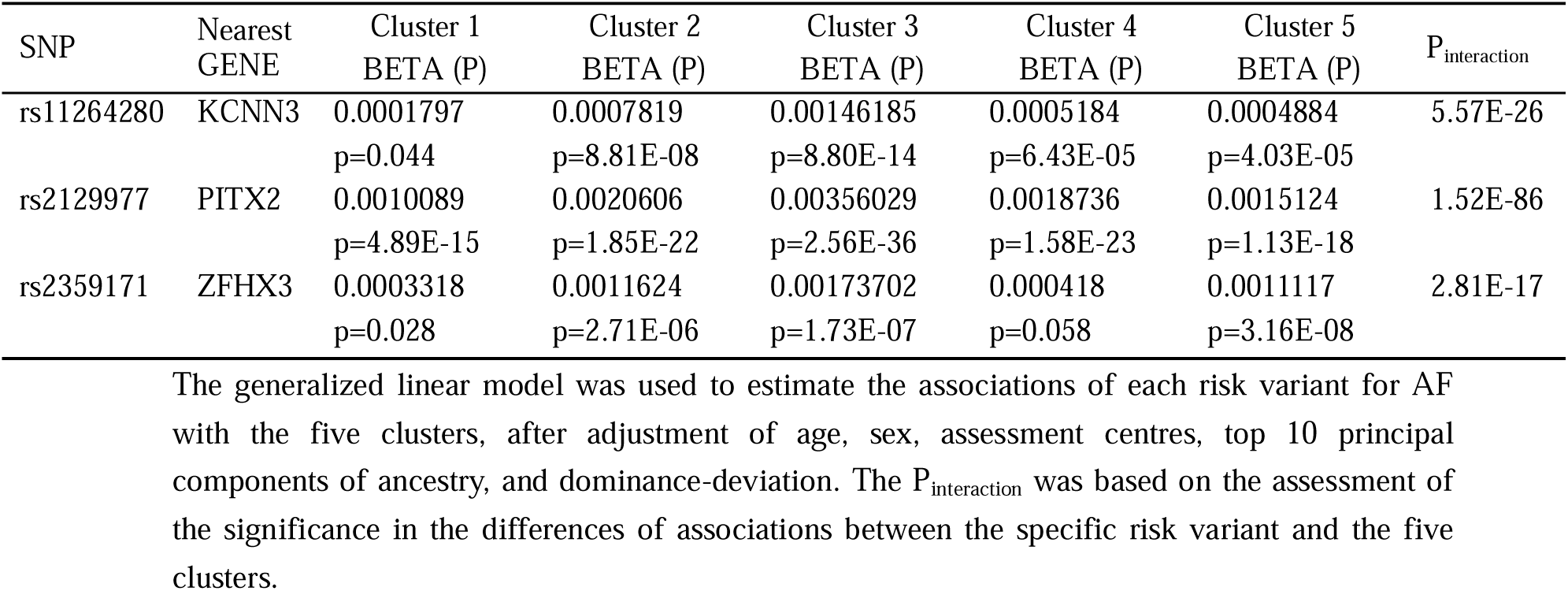
Genetic associations of risk variants for AF reaching significance of genome-wide association study with at least one clusters.

### Practicality of the clustering algorithm and replication in a new cohort

The characteristics were similar between original clusters in main analysis and re-clusters that used the shortest Pearson distance from the clustering center (**Supplementary Figure S6 and Table S14**). Then, we clustered participants with newly-onset AF in 2 years after baseline and obtained similar five clusters to the original clusters (**Supplementary Figure S7)**. In addition, we found that the difference in disease risks were similar to those in main analyses (**Supplementary Table S15**).

## Discussion

Our findings provided a new clustering of AF patients, which might help to identify patients at high risk for AF related complications and guide the choice of treatment. We found that AF patients could be classified into five clusters of risk profiles. Cluster 1 (acute illness-related AF) was characterized by young age and acute illnesses, cluster 2 (mental health-related AF) by poor mental health, cluster 3 (CVD-related AF) by a high prevalence of cardiovascular diseases and acute illnesses, cluster 4 (immune-and-renal disease related AF) by high prevalence of immune disease and renal dysfunction and cluster 5 (respiratory-and-metabolic disease-related AF) by prominent respiratory and metabolic problems. In addition to population characteristics, the risk of death and important complications, as well as loss of life expectancy, differed between clusters. The underlying genetic background behind the five clusters also varied. Furthermore, we applied this cluster algorithm based on the existing model to classify AF patients in clinical setting.

Among five clusters, cluster 5 (respiratory- and metabolic disease-related AF) represented the most severe type of AF, and this cohort deserved more attention, intensified treatment, and long-term follow-up to reduce the risk of AF complications. Patients in cluster 5 had a significantly higher risk of death and reduction of life expectancy than the other clusters, which underscored the fact that metabolic and respiratory co-morbidities should be prioritized in managing AF patients. On one hand, COPD and apnea, as two respiratory factors strongly associated with AF, both contribute to hypoxemia. Patients with underlying cardiac disease including AF have a higher risk of death and AMI under the condition of prolonged hypoxemia ^23^. On the other hand, many metabolic risk factors contribute to the development of AF ^24^, leaded by overweight/obesity^12^. We identified that ZFHX3 (rs2359171) was only associated with cluster 5. ZFHX3 was reported in genome-wide association analyses of both BMI and AF ^22,25^, which may be the underlying genetic shared between overweight/obesity and AF. Furthermore, patients in cluster 5 also had highest risk of HF. Anticoagulation to prevent stroke has already been integrated part of Atrial fibrillation Better Care (ABC) holistic pathway ^3^. Primary prevention of HF in AF seems to be less considered. We suppose that in patients with respiratory- and metabolic disease-related AF, use of sodium/glucose cotransporter 2 inhibitor might improve HF outcome, which warrants further trials.

Notably, we found that individuals in cluster 2 (mental health-related AF) lost the second most life expectancy, out of the five clusters. Cluster 2 had a unique profile, showing the highest prevalence of psychological problems (loneliness and depression) among five clusters. Although psychological problems are not included in the guidelines as risk factors for AF, the role of mental health especially depression in the pathogenesis of AF has been acknowledged, ^26,27^. Our results provided further evidence that KCNN3 (rs2129977) was associated with mental health-related AF. KCNN3, performing the function of promoting action potential repolarization^28^, was proved to be associated with the development of AF ^22,29^ and influenced by depressive illness ^30^, which indicates a potential mechanism by which mood influences the onset of AF. Mental health deserves more attention in the prevention and control of AF. Further, the prevention of CIS should be attached more importance in those patients, considering the second higher risk of CIS among the five clusters.

We found that cluster 1 (acute illness-related AF) was the cluster with the best prognosis among all clusters. This group clusters 45% people younger than 55 years old in this study, suggesting the importance of acute illness on development of AF in young and middle-aged patients. Further, acute illness (major operation, pneumonia) and immune-related diseases are both responsible for inflammation and oxidative stress, and eventually induces AF ^31–33^. Therefore, we believe it’s worthwhile to control immune response and rationally use antioxidants in these patients. Of note, an increasing number of randomized controlled trials have demonstrated the significant effect of antioxidants and control of the immune response in the prevention of AF ^34,35^.

Traditional AF classification is based on presentation, duration and spontaneous termination of AF episodes. However, in clinical practice, a large proportion of patients presented insidious symptoms^36^, which makes it difficult to be classified by AF episodes. In contrast, the information needed for our classification method is more easily collected and patients with unspecified AF coded by ICD were further classified into specific clusters.

In the present study, we provided a feasible score of 1 to 5 for patients, classified into clusters 1 to 5 in order, in terms of clinical assessment in Substrate severity of AF. Due to the underlying heterogeneity of the influence of different risk factors on AF, it is not advisable to simply list the number of risk factors in a patient as a risk score. We took the diversity of AF comorbidities and the heterogeneity of their prognostic impact into account. By integrating much available risk factors as possible into the clustering analysis, our classification method can refine AF subtyping and guide more accurate risk stratification.

Our study has several limitations. First, we cannot claim that this is the best clustering for AF although we made full use of risk factors from previous evidence. Moreover, whether there is stability of clustering for patients with distant centers of clustering needs to be confirmed in future prospective cohorts. Second, the main applicable ethnicity for our clustering algorithm is European and the stability of this algorithm should be further validated in other cohorts. Third, the generalizability of our findings was limited to population older than 40 because of the composition of the participants in UKB. Further research is warranted in the population presenting early-onset AF.

### Clinical implication

The identification of clusters based on demographic, anthropometric, and clinical characteristics allows for a more personalized approach to patient care and interventions based on specific risk profiles. The recognition of different AF clusters (“acute illness-related AF”, “mental health-related AF”, “CVD-related AF”, “immune-and-renal disease related AF”, and “respiratory-and-metabolic disease-related AF”) can help to guide physicians to understand the diverse pathophysiological mechanisms underlying AF. Strategies of therapy targeting the specific characteristics of each cluster may lead to more effective prevention of complications and outcome improvement. Furthermore, the observation of risk of death and major complications in patients within each AF cluster underscores the importance of risk stratification and long-term monitoring. Patients in clusters with high-risk profiles, such as “respiratory- and metabolic disease-related AF”, may require the most positive management to mitigate adverse outcomes.

## Conclusion

Different AF subtypes exhibited unique features and difference in mortality and complications. By integrating health habits, demographic characteristics, biochemical indicators, and disease diagnostic information, we provide clinically valuable clustering in patients with AF, gives the opportunity to benefit from precision interventions.

## Data Availability

The data underlying this article are available in UK Biobank, at https://www.ukbiobank.ac.uk/.

https://ukbiobank.dnanexus.com/

## Ethnic declaration

The UKB received ethical approval from the UK National Health Service, National Research Ethics Service North West, the National Information Governance Board for Health and Social Care in England and Wales, and the Community Health Index Advisory Group in Scotland. All procedures followed were in accordance with the ethical standards of the responsible committee on human experimentation (institutional and national) and with the Helsinki Declaration of 1975, as revised in 2008. All participants provided written informed consent. This study was approved by the UK Biobank (application number 77740).

## Data availability statement

The data underlying this article are available in UK Biobank, at https://www.ukbiobank.ac.uk/. The datasets were derived from sources in the public domain: https://ukbiobank.dnanexus.com/.

## Code availability

The R “clustercnp” package for clustering AF patients is available on https://github.com/JiangLi-1/clustercnp.

## Acknowledgements

This research has been conducted using the UK Biobank Resource under Application Number 77740. This study was supported by Shanghai Municipal Health Commission (2022XD017), Clinical Research Plan of SHDC (SHDC2020CR4006), National Natural Science Foundation of China (82170870), Shanghai Municipal Human Resources and Social Security Bureau (2020074), Innovative Research Team of High-level Local Universities in Shanghai (SHSMU-ZDCX20212501). The funders played no role in the design or conduction of the study; in the collection, management, analysis, or interpretation of the data; or in the preparation, review, or approval of the article.

## Declarations

No potential conflicts of interest relevant to this article were reported.

## Author contributions

N.W. and Y.L. concepted this paper. Y.Y., J.L. and N.W. wrote the manuscript, researched data, and reviewed/edited the manuscript. N.W. and X.T. reviewed/edited the manuscript. J.L., Y.S., H.Z., Y.W. and B.W. contributed to data acquisition and revised it critically for important intellectual content. All authors approved the final manuscript.

