## Supplementary materials for "Outcome and life expectancy associated with novel subgroups of atrial fibrillation: a data-driven cluster analysis"

Supplementary Table S1. Definitions and sources of information for outcomes in UK Biobank

Supplementary Table S2. Definition of risk factors

Supplementary Table S3. Estimate of each risk factor in LASSO model

Supplementary Table S4. Scores of 6 categories

Supplementary Table S5. Incidences of death and complications during follow-up

Supplementary Table S6. Basic adjusted hazard ratios of death associated with AF cluster

Supplementary Table S7. Basic adjusted hazard ratios of complications associated with AF cluster

Supplementary Table S8. Additional adjusted hazard ratios of complications associated with AF cluster

Supplementary Table S9. Reductions of life expectency for participants in AF cluster 2 to 4, comparing with cluster 1

Supplementary Table S10. Redistribution of types of AF under the ICD classification into new clusters

Supplementary Table S11. Hazard ratios of death and major complications for the five AF cluster, comparing with non-AF

Supplementary Table S12. Reductions of life expectency for participants in the five AF clusters comparing with non-AF participants

Supplementary Table S13. Genetic associations of risk variants for AF with the five clusters Supplementary Table S14. The difference in participant number between origin cluster and re-cluster

Supplementary Table S15. Hazard ratios of death and complications associated with AF cluster in participants with newly-onset AF in 2 years after baseline.

Supplementary Method 1. LASSO algorithm

Supplementary Method 2. Factor analysis protocol

Supplementary Method 3. Consensus clustering algorithm

Supplementary Figure S1. Study design

Supplementary Figure S2. Number of variables included in models for different values of lambda and the estimates of corresponding variables

Supplementary Figure S3. The C-index and the partial likelihood deviation of the models for different values of lambda.

Supplementary Figure S4. Pairwise comparisons of the cluster feature variables. Supplementary Figure S5. Reductions of life expectency for participants in the five AF clusters comparing with non-AF participants.

Supplementary Figure S6. Cluster features differed among five clusters in re-clustering analysis among the participants with AF before baseline.

Supplementary Figure S7. Cluster features differed among five clusters under new algorithm among the participants with newly-onset AF within 2 years after baseline.

Extended Figure 1. The rubble plot of eigen value based on different numbers of factor. Extended Figure 2. The factor loading of 16 risk factors in 6 factors.

Extended Figure 3. (A) The cumulative distribution function (CDF) for each cluster number (K). (B) Relative change in area under the CDF curve comparing K and K – 1.

Extended Figure 4. Consensus matrix of probability of each participant categorized into the same cluster with other participants.

Supplementary Table S1. Definitions and sources of information for outcomes in UK Biobank.

| Cardiovascular conditions | Number of case | Field ID | ICD-10 diagnosis (first occurrences, category 2409) | Other |
| --- | --- | --- | --- | --- |
| Cerebral Ischemic stroke | 630 | 41270 | I63 |  |
| Acute myocardial infarction | 1364 | 41270 | I21 |  |
| Heart failure | 2058 | 41270 | I50 | I11.0, I13.0, I13.2 |
| All cause death | 1462 | 40000 | / |  |
| cardiovascular death | 572 | 40000  40001 | I00-I99 |  |
| ICD, International Classification of Disease. | |  |  |  |

Supplementary Table S2. Definition of risk factors

|  | Risk factors | Definition | Field ID |
| --- | --- | --- | --- |
| Demographic factors | Age | Age on baseline. It was based on date of birth and date of attending an initial assessment centre. | 21022 |
|  | Male sex | Sex as determined from genotyping analysis. | 22001 |
|  | Caucasian | Participants who were classified as Caucasian in genetic ethnic group. | 22006 |
|  | Less education | Less than high school. | 6138 |
|  | Townsend Deprivation Index (TDI) | Townsend deprivation index ≥ 1.34 (Highest quintile of Townsend deprivation index of all individuals) | 738 |
|  | Severe air pollution | Nitrogen dioxide (NO2) ≥ 32.49 macro-g/m3 (highest quintile of NO2 of all individuals) or nitrogen oxides (NOx) ≥ 53.22 macro-g/m3 (highest quintile of NOx of all individuals) | 24003, 24004 |
| Health behaviours | Current smoking | Smoking tobacco most or all days | 1239 |
|  | Excessive alcohol intake | Alcohol intake ≥ 24g/d | 1568, 1578, 1588, 1598, 1608, 4407, 4418, 4429, 4440 |
|  | Physical inactivity | <150 minutes/week of moderate intensity, <75 minutes/week of vigorous intensity, or an equivalent combination | 894, 884,  914, 904 |
|  | Excessive physical activity | >55 MET-hours/week (>3300 MET-minutes/week) | 22040 |
|  | Loneliness | Participant answered “YES” in “Do you often feel lonely?” | 2020 |
|  | Broad depression | Participant answered “YES” in “Have you ever seen a psychiatrist for nerves, anxiety, tension or depression?” | 2100 |
| Clinical comorbidities | Hypertension | Participants who were diagnosed with hypertension, or regularly took medications for blood pressure at baseline, or systolic blood pressure ≥140 mmHg, or diastolic blood pressure ≥90mmHg | 6177, 6153 |
|  |  |  | 93,4080 |
|  |  |  | 94,4079 |
|  | Low Low-density lipoprotein cholesterol (LDL-C) | Low-density lipoprotein cholesterol < 2.81 mmol/L (Lowest quintile) | 30780 |
|  | Low triglycerides | Triglycerides < 0.965 mmol/L (Lowest quintile) | 30870 |
|  | Diabetes mellitus/ prediabetes | Participants who were diagnosed with diabetes or took medications for diabetes at baseline, or HbA1c ≥ 5.7% | 6177, 6153, 30750, 30740 |
|  | Overweight/obesity | BMI ≥25 kg/m^2^ | 21001 |
|  | Elevated C-reactive protein (CRP) | CRP ≥3.12mg/L (Highest quintile of CRP of all individuals) | 30710 |
|  | Sleep apnoea | Sleep apnoea at baseline | ICD-10 (G47.3) |
|  | Chronic obstructive pulmonary disease (COPD) | Chronic obstructive pulmonary disease at baseline | ICD-10 (J42-J44) |
|  | Renal dysfunction | Chronic renal disease at baseline, or eGFR <60mL*min^-1^ *(1.73 m^2^)^-1^ | ICD-10 (N18)  30700, 30720 |
|  | Cardiovascular conditions | Heart failure at baseline | ICD-10 (I50, I11.0, I13.0, I13.2) |
|  |  | Valvular disease at baseline | ICD-10 (I05, I06, I07, I08, I34, I35, I36, I37) |
|  |  | Coronary heart disease at baseline | ICD-10 (I21, I22, I23, I24, I25.1, I25.2, I25.5, I25.6, I25.8, I25.9) |
|  |  | Congenital heart disease at baseline | ICD-10 (Q20, Q21, Q22, Q23, Q24, Q25) |
|  |  | Sick sinus syndrome at baseline | I49.5 |
|  |  | Wolff-Parkinson White at baseline | 20002 (coding 1484) |
|  | Immune mediated disease | Hypo- and hyperthyroidism at baseline | ICD-10 (E02, E03, E05) |
|  |  | Coeliac disease, rheumatoid arthritis, and psoriasis at baseline | ICD-10 (K90.0, M05, M06, L40) |
|  | Acute illness | History of sepsis at baseline | ICD-10 (A02.1, A22.7, A32.7, A40, A41, A42.7, B37.7, O85, R65.1, R57.2) |
|  |  | History of major operations at baseline | 2415, 2844, 136 |
|  |  | Pneumonia at baseline | ICD-10 (J12, J13, J14, J15, J16, J17, J18) |
| Genetic factor | High genetic risk | Genetic risk score (GRS) > 6.72  (Highest quintile of GRS of all individuals) | GWAS data |

Supplementary Table S3. Estimate of each risk factor in LASSO model.

| Risk factors | Estimates |
| --- | --- |
| Less education | 0.04750041 |
| Socioeconomic deprivation | 0.11090754 |
| Severe air pollution | 0.02156172 |
| Current smoking | 0.12904952 |
| Excessive alcohol intake | 0.10829389 |
| Physical inactivity | . |
| Excessive physical activity | . |
| Loneliness | 0.07252611 |
| Broad depression | 0.08311185 |
| High blood pressure | 0.28957775 |
| Low LDL-C | 0.13400976 |
| Low triglycerides | 0.13570686 |
| Diabetes mellitus/prediabetes | 0.05741334 |
| Obesity | 0.20803518 |
| Elevated CRP | 0.19983445 |
| Sleep apnoea | 0.33999023 |
| COPD | 0.35530800 |
| Renal dysfunction | 0.40395812 |
| Cardiovascular diseases | 0.47051097 |
| Immune mediated disease | 0.05022985 |
| Acute illness | 0.16521931 |
| Male sex | 0.57508578 |
| Caucasian race | 0.04914593 |
| Age | 0.09504056 |
| High genetic risk | 0.74253318 |

COPD, chronic obstructive pulmonary disease; LDL-C, low-density lipoprotein cholesterol; CRP, C-reactive protein.

Supplementary Table S4. Scores of 6 categories

| Categories | Variables | Score range |
| --- | --- | --- |
| Metabolic factors | Hypertension, overweight/obesity,  diabetes mellitus/prediabetes | 0-3 |
| Respiratory factors | COPD, sleep apnoea, current smoking and elevated CRP | 0-4 |
| Cardiovascular factors | Cardiovascular diseases (heart failure, valvular disease, coronary heart disease, congenital heart disease), low LDL-C, low triglycerides | 0-6 |
| Renal and immunity diseases | Renal dysfunction, immune mediated disease, excessive alcohol intake | 0-3 |
| Mental health | Loneliness, broad depression | 0-2 |
| Acute illness | Major operations, sepsis, pneumonia | 0-3 |

Supplementary Table S5. Incidences of death and complications during follow-up.

|  | Cluster 1 | Cluster 2 | Cluster 3 | Cluster 4 | Cluster 5 |
| --- | --- | --- | --- | --- | --- |
| All-cause death | 61 (10.4%) | 243 (15.3%) | 284 (9.8%) | 305 (24.3%) | 569 (54.4%) |
| Cardiovascular death | 44 (7.5%) | 168 (10.5%) | 173 (5.9%) | 197 (15.7%) | 384 (36.7%) |
| Acute myocardial infarction | 66 (11.2%) | 197 (12.4%) | 278 (9.5%) | 225 (18%) | 598 (57.2%) |
| Heart failure | 138 (23.5%) | 336 (21.1%) | 415 (14.3%) | 364 (29.1%) | 805 (77%) |
| Cerebral Ischemic stroke | 34 (5.8%) | 101 (6.3%) | 138 (4.7%) | 109 (8.7%) | 248 (23.7%) |

Supplementary Table S6. Basic adjusted hazard ratios of death associated with AF cluster.

|  | HR (95%CI) for all-cause death | | | | |
| --- | --- | --- | --- | --- | --- |
| Reference | Cluster 1 | Cluster 2 | Cluster 3 | Cluster 4 | Cluster 5 |
| Cluster 1 | 1 | 1.85(1.4,2.43) | 1.95(1.49,2.53) | 1.95(1.47,2.58) | 3.21(2.44,4.23) |
| Cluster 2 | 0.54(0.41,0.71) | 1 | 1.05(0.91,1.22) | 1.06(0.89,1.25) | 1.74(1.48,2.05) |
| Cluster 3 | 0.51(0.39,0.67) | 0.95(0.82,1.09) | 1 | 1(0.86,1.16) | 1.65(1.44,1.9) |
| Cluster 4 | 0.51(0.39,0.68) | 0.95(0.8,1.12) | 1(0.86,1.16) | 1 | 1.65(1.39,1.95) |
| Cluster 5 | 0.31(0.24,0.41) | 0.57(0.49,0.67) | 0.61(0.53,0.7) | 0.61(0.51,0.72) | 1 |
|  | HR (95%CI) for cardiovascular death | | | | |
| Reference | Cluster 1 | Cluster 2 | Cluster 3 | Cluster 4 | Cluster 5 |
| Cluster 1 | 1 | 1.57(1.13,2.19) | 1.83(1.34,2.5) | 1.88(1.35,2.61) | 2.9(2.09,4.03) |
| Cluster 2 | 0.64(0.46,0.89) | 1 | 1.17(0.97,1.4) | 1.19(0.97,1.48) | 1.85(1.51,2.27) |
| Cluster 3 | 0.55(0.4,0.75) | 0.86(0.72,1.03) | 1 | 1.02(0.85,1.23) | 1.59(1.34,1.88) |
| Cluster 4 | 0.53(0.38,0.74) | 0.84(0.68,1.04) | 0.98(0.81,1.17) | 1 | 1.55(1.26,1.9) |
| Cluster 5 | 0.34(0.25,0.48) | 0.54(0.44,0.66) | 0.63(0.53,0.75) | 0.65(0.53,0.79) | 1 |

Cox proportional hazards models were adjusted for sex and race.

Supplementary Table S7. Basic adjusted hazard ratios of complications associated with AF cluster.

|  | HR (95%CI) for AMI | | | | |
| --- | --- | --- | --- | --- | --- |
| Reference | Cluster 1 | Cluster 2 | Cluster 3 | Cluster 4 | Cluster 5 |
| Cluster 1 | 1 | 1.75(1.34,2.29) | 1.97(1.53,2.55) | 1.45(1.09,1.91) | 2.17(1.65,2.86) |
| Cluster 2 | 0.57(0.44,0.75) | 1 | 1.13(0.98,1.3) | 0.83(0.69,0.99) | 1.24(1.04,1.48) |
| Cluster 3 | 0.51(0.39,0.65) | 0.88(0.77,1.02) | 1 | 0.73(0.62,0.86) | 1.1(0.94,1.28) |
| Cluster 4 | 0.69(0.52,0.91) | 1.21(1.01,1.45) | 1.37(1.16,1.6) | 1 | 1.5(1.24,1.82) |
| Cluster 5 | 0.46(0.35,0.61) | 0.8(0.67,0.96) | 0.91(0.78,1.06) | 0.67(0.55,0.81) | 1 |
|  | HR (95%CI) for HF | | | | |
| Reference | Cluster 1 | Cluster 2 | Cluster 3 | Cluster 4 | Cluster 5 |
| Cluster 1 | 1 | 1.17(0.97,1.42) | 1.23(1.03,1.48) | 1.19(0.98,1.46) | 1.67(1.37,2.03) |
| Cluster 2 | 0.85(0.7,1.03) | 1 | 1.05(0.93,1.18) | 1.02(0.88,1.18) | 1.42(1.24,1.64) |
| Cluster 3 | 0.81(0.68,0.97) | 0.95(0.85,1.07) | 1 | 0.97(0.85,1.1) | 1.35(1.2,1.53) |
| Cluster 4 | 0.84(0.69,1.02) | 0.98(0.85,1.13) | 1.03(0.91,1.17) | 1 | 1.4(1.21,1.62) |
| Cluster 5 | 0.6(0.49,0.73) | 0.7(0.61,0.81) | 0.74(0.65,0.84) | 0.72(0.62,0.83) | 1 |
|  | HR (95%CI) for CIS | | | | |
| Reference | Cluster 1 | Cluster 2 | Cluster 3 | Cluster 4 | Cluster 5 |
| Cluster 1 | 1 | 1.56(1.07,2.27) | 1.54(1.08,2.21) | 1.45(0.99,2.14) | 2(1.36,2.95) |
| Cluster 2 | 0.64(0.44,0.94) | 1 | 0.99(0.8,1.22) | 0.93(0.72,1.21) | 1.29(1,1.65) |
| Cluster 3 | 0.65(0.45,0.93) | 1.01(0.82,1.24) | 1 | 0.94(0.75,1.19) | 1.3(1.04,1.63) |
| Cluster 4 | 0.69(0.47,1.02) | 1.07(0.83,1.39) | 1.06(0.84,1.34) | 1 | 1.38(1.05,1.81) |
| Cluster 5 | 0.5(0.34,0.73) | 0.78(0.6,1) | 0.77(0.61,0.96) | 0.73(0.55,0.95) | 1 |

Cox proportional hazards models were adjusted for sex and race. AMI, acute myocardial infarction, HF, heart failure, CIS, cerebral ischemic stroke.

Supplementary Table S8. Additional adjusted hazard ratios of complications associated with AF cluster.

|  | HR (95%CI) for all-cause death | | | | |
| --- | --- | --- | --- | --- | --- |
| Reference | Cluster 1 | Cluster 2 | Cluster 3 | Cluster 4 | Cluster 5 |
| Cluster 1 | 1 | 1.66(1.25,2.21) | 1.82(1.38,2.4) | 1.85(1.38,2.47) | 2.87(2.16,3.82) |
| Cluster 2 | 0.6(0.45,0.8) | 1 | 1.1(0.95,1.27) | 1.11(0.94,1.33) | 1.73(1.47,2.04) |
| Cluster 3 | 0.55(0.42,0.72) | 0.91(0.79,1.05) | 1 | 1.01(0.87,1.18) | 1.58(1.37,1.82) |
| Cluster 4 | 0.54(0.4,0.72) | 0.9(0.75,1.07) | 0.99(0.85,1.15) | 1 | 1.55(1.31,1.84) |
| Cluster 5 | 0.35(0.26,0.46) | 0.58(0.49,0.68) | 0.63(0.55,0.73) | 0.64(0.54,0.76) | 1 |
|  | HR (95%CI) for cardiovascular death | | | | |
| Reference | Cluster 1 | Cluster 2 | Cluster 3 | Cluster 4 | Cluster 5 |
| Cluster 1 | 1 | 1.35(0.96,1.9) | 1.65(1.19,2.28) | 1.71(1.21,2.42) | 2.46(1.75,3.46) |
| Cluster 2 | 0.74(0.53,1.04) | 1 | 1.22(1.01,1.46) | 1.27(1.02,1.57) | 1.82(1.48,2.24) |
| Cluster 3 | 0.61(0.44,0.84) | 0.82(0.68,0.99) | 1 | 1.04(0.87,1.25) | 1.5(1.26,1.78) |
| Cluster 4 | 0.58(0.41,0.82) | 0.79(0.64,0.98) | 0.96(0.8,1.15) | 1 | 1.44(1.17,1.77) |
| Cluster 5 | 0.41(0.29,0.57) | 0.55(0.45,0.67) | 0.67(0.56,0.8) | 0.7(0.56,0.86) | 1 |

|  | HR (95%CI) for AMI | | | | |
| --- | --- | --- | --- | --- | --- |
| Reference | Cluster 1 | Cluster 2 | Cluster 3 | Cluster 4 | Cluster 5 |
| Cluster 1 | 1 | 1.25(0.96,1.65) | 1.38(1.07,1.79) | 1.07(0.81,1.42) | 1.46(1.1,1.93) |
| Cluster 2 | 0.8(0.61,1.05) | 1 | 1.1(0.95,1.27) | 0.85(0.71,1.03) | 1.16(0.97,1.39) |
| Cluster 3 | 0.73(0.56,0.94) | 0.91(0.79,1.05) | 1 | 0.77(0.66,0.91) | 1.06(0.91,1.24) |
| Cluster 4 | 0.94(0.71,1.24) | 1.17(0.98,1.41) | 1.29(1.1,1.52) | 1 | 1.37(1.13,1.66) |
| Cluster 5 | 0.69(0.52,0.91) | 0.86(0.72,1.03) | 0.94(0.81,1.1) | 0.73(0.6,0.89) | 1 |
|  | HR (95%CI) for HF | | | | |
| Reference | Cluster 1 | Cluster 2 | Cluster 3 | Cluster 4 | Cluster 5 |
| Cluster 1 | 1 | 1(0.82,1.22) | 1.07(0.89,1.29) | 1.05(0.86,1.28) | 1.39(1.14,1.7) |
| Cluster 2 | 1(0.82,1.22) | 1 | 1.07(0.95,1.21) | 1.05(0.91,1.22) | 1.39(1.2,1.6) |
| Cluster 3 | 0.94(0.78,1.13) | 0.94(0.83,1.06) | 1 | 0.98(0.86,1.12) | 1.3(1.15,1.47) |
| Cluster 4 | 0.95(0.78,1.17) | 0.95(0.82,1.1) | 1.02(0.9,1.16) | 1 | 1.32(1.14,1.54) |
| Cluster 5 | 0.72(0.59,0.88) | 0.72(0.62,0.83) | 0.77(0.68,0.87) | 0.76(0.65,0.88) | 1 |
|  | HR (95%CI) for CIS | | | | |
| Reference | Cluster 1 | Cluster 2 | Cluster 3 | Cluster 4 | Cluster 5 |
| Cluster 1 | 1 | 1.34(0.92,1.97) | 1.31(0.9,1.88) | 1.27(0.86,1.89) | 1.68(1.14,2.49) |
| Cluster 2 | 0.74(0.51,1.09) | 1 | 0.97(0.79,1.2) | 0.95(0.73,1.23) | 1.25(0.97,1.61) |
| Cluster 3 | 0.77(0.53,1.11) | 1.03(0.83,1.27) | 1 | 0.98(0.77,1.23) | 1.29(1.03,1.62) |
| Cluster 4 | 0.79(0.53,1.17) | 1.06(0.81,1.37) | 1.03(0.81,1.29) | 1 | 1.32(1.01,1.73) |
| Cluster 5 | 0.59(0.4,0.88) | 0.8(0.62,1.03) | 0.78(0.62,0.97) | 0.76(0.58,0.99) | 1 |

Cox proportional hazards models were adjusted for sex, race, GRS, education, TDI, and air pollution. AMI, acute myocardial infarction, HF, heart failure, CIS, cerebral ischemic stroke.

Supplemental Table S9. Reductions of life expectency for participants in AF cluster 2 to 4, comparing with cluster 1.

| Age | Cluster 2 | Cluster 3 | Cluster 4 | Cluster 5 |
| --- | --- | --- | --- | --- |
| 40 | 30.4(28.95,31.88) | 12.11(11.14,13.1) | 8.73(7.91,9.56) | 32.62(31.12,34.15) |
| 41 | 29.42(27.95,30.92) | 12.04(11.09,13.01) | 8.83(8.02,9.64) | 31.67(30.14,33.23) |
| 42 | 28.47(27,30) | 11.94(11.02,12.89) | 8.89(8.11,9.68) | 30.74(29.2,32.33) |
| 43 | 27.63(26.14,29.16) | 11.85(10.94,12.77) | 8.93(8.17,9.7) | 29.91(28.35,31.52) |
| 44 | 26.79(25.29,28.34) | 11.73(10.84,12.64) | 8.95(8.2,9.7) | 29.08(27.51,30.71) |
| 45 | 26.19(24.69,27.74) | 11.64(10.77,12.53) | 8.95(8.22,9.69) | 28.48(26.9,30.12) |
| 46 | 25.74(24.25,27.3) | 11.56(10.71,12.44) | 8.94(8.23,9.67) | 28.04(26.46,29.68) |
| 47 | 25.22(23.73,26.77) | 11.47(10.63,12.33) | 8.93(8.23,9.64) | 27.52(25.93,29.16) |
| 48 | 24.76(23.28,26.31) | 11.37(10.55,12.22) | 8.9(8.22,9.6) | 27.05(25.47,28.7) |
| 49 | 24(22.52,25.56) | 11.21(10.4,12.04) | 8.86(8.18,9.54) | 26.29(24.7,27.95) |
| 50 | 22.41(20.91,24) | 10.86(10.08,11.67) | 8.74(8.09,9.41) | 24.68(23.04,26.39) |
| 51 | 21.41(19.91,23.01) | 10.63(9.87,11.42) | 8.66(8.02,9.3) | 23.66(22.01,25.39) |
| 52 | 20.91(19.41,22.5) | 10.51(9.77,11.28) | 8.61(7.99,9.24) | 23.14(21.51,24.88) |
| 53 | 20.16(18.67,21.75) | 10.33(9.6,11.08) | 8.53(7.92,9.15) | 22.37(20.74,24.12) |
| 54 | 19.77(18.3,21.35) | 10.23(9.51,10.97) | 8.49(7.89,9.1) | 21.98(20.35,23.72) |
| 55 | 19.13(17.67,20.7) | 10.05(9.35,10.78) | 8.4(7.82,9) | 21.31(19.69,23.05) |
| 56 | 18.67(17.22,20.24) | 9.92(9.24,10.63) | 8.34(7.77,8.92) | 20.83(19.21,22.57) |
| 57 | 18.05(16.61,19.6) | 9.74(9.07,10.44) | 8.24(7.69,8.82) | 20.18(18.57,21.92) |
| 58 | 17.73(16.31,19.28) | 9.65(8.99,10.33) | 8.19(7.64,8.76) | 19.85(18.25,21.59) |
| 59 | 17.42(16.01,18.96) | 9.55(8.91,10.23) | 8.14(7.6,8.69) | 19.53(17.93,21.26) |
| 60 | 16.83(15.43,18.36) | 9.36(8.73,10.02) | 8.03(7.5,8.57) | 18.9(17.31,20.64) |
| 61 | 16.43(15.05,17.95) | 9.23(8.61,9.88) | 7.96(7.44,8.49) | 18.48(16.9,20.22) |
| 62 | 16.08(14.71,17.58) | 9.11(8.5,9.74) | 7.88(7.38,8.4) | 18.11(16.54,19.83) |
| 63 | 15.68(14.33,17.18) | 8.97(8.38,9.59) | 7.8(7.3,8.31) | 17.69(16.13,19.41) |
| 64 | 15.28(13.94,16.77) | 8.83(8.25,9.44) | 7.71(7.22,8.21) | 17.26(15.71,18.98) |
| 65 | 14.79(13.47,16.27) | 8.65(8.08,9.24) | 7.59(7.12,8.08) | 16.74(15.19,18.46) |
| 66 | 14.45(13.14,15.92) | 8.52(7.96,9.1) | 7.51(7.04,7.99) | 16.38(14.84,18.09) |
| 67 | 14.12(12.82,15.58) | 8.39(7.84,8.96) | 7.42(6.97,7.89) | 16.02(14.49,17.74) |
| 68 | 13.78(12.49,15.22) | 8.25(7.71,8.81) | 7.33(6.88,7.79) | 15.65(14.13,17.37) |
| 69 | 13.44(12.17,14.88) | 8.11(7.58,8.65) | 7.23(6.79,7.68) | 15.29(13.78,17) |
| 70 | 13.07(11.82,14.49) | 7.95(7.44,8.48) | 7.12(6.69,7.56) | 14.89(13.38,16.6) |
| 71 | 12.7(11.46,14.11) | 7.78(7.28,8.31) | 7(6.59,7.43) | 14.49(12.99,16.2) |
| 72 | 12.33(11.1,13.73) | 7.61(7.13,8.13) | 6.88(6.48,7.3) | 14.09(12.6,15.81) |
| 73 | 11.95(10.75,13.35) | 7.44(6.97,7.94) | 6.75(6.36,7.16) | 13.68(12.2,15.41) |
| 74 | 11.59(10.39,12.98) | 7.27(6.8,7.76) | 6.62(6.24,7.02) | 13.28(11.81,15.01) |
| 75 | 11.2(10.02,12.58) | 7.08(6.63,7.55) | 6.48(6.1,6.87) | 12.86(11.39,14.59) |
| 76 | 10.79(9.63,12.16) | 6.87(6.43,7.34) | 6.32(5.95,6.7) | 12.42(10.96,14.16) |
| 77 | 10.38(9.23,11.75) | 6.66(6.23,7.11) | 6.15(5.8,6.52) | 11.97(10.51,13.72) |
| 78 | 9.96(8.83,11.32) | 6.44(6.03,6.88) | 5.98(5.63,6.33) | 11.51(10.06,13.28) |
| 79 | 9.52(8.41,10.88) | 6.2(5.8,6.63) | 5.79(5.45,6.13) | 11.03(9.59,12.81) |
| 80 | 9.06(7.96,10.41) | 5.95(5.56,6.36) | 5.58(5.26,5.91) | 10.52(9.09,12.33) |
| 81 | 8.6(7.51,9.93) | 5.68(5.31,6.08) | 5.36(5.05,5.68) | 10.01(8.58,11.84) |
| 82 | 8.13(7.07,9.46) | 5.41(5.05,5.8) | 5.13(4.83,5.45) | 9.5(8.07,11.35) |
| 83 | 7.65(6.61,8.97) | 5.13(4.78,5.5) | 4.89(4.6,5.19) | 8.96(7.55,10.84) |
| 84 | 7.16(6.15,8.48) | 4.84(4.51,5.2) | 4.64(4.36,4.93) | 8.42(7.02,10.34) |
| 85 | 6.66(5.67,7.97) | 4.53(4.21,4.87) | 4.37(4.11,4.64) | 7.87(6.47,9.82) |
| 86 | 6.15(5.19,7.45) | 4.21(3.91,4.54) | 4.09(3.84,4.35) | 7.3(5.92,9.3) |
| 87 | 5.63(4.7,6.91) | 3.88(3.59,4.19) | 3.79(3.55,4.04) | 6.71(5.35,8.76) |
| 88 | 5.08(4.19,6.35) | 3.53(3.26,3.82) | 3.47(3.25,3.7) | 6.1(4.77,8.21) |
| 89 | 4.55(3.7,5.79) | 3.17(2.93,3.45) | 3.14(2.93,3.36) | 5.49(4.19,7.66) |
| 90 | 4(3.2,5.22) | 2.81(2.58,3.07) | 2.8(2.61,3.01) | 4.86(3.62,7.12) |
| 91 | 3.48(2.74,4.67) | 2.46(2.25,2.69) | 2.47(2.3,2.66) | 4.27(3.08,6.61) |
| 92 | 2.96(2.27,4.1) | 2.1(1.91,2.31) | 2.13(1.97,2.3) | 3.65(2.54,6.09) |
| 93 | 2.45(1.84,3.53) | 1.75(1.58,1.93) | 1.78(1.65,1.94) | 3.05(2.03,5.61) |
| 94 | 1.94(1.42,2.95) | 1.4(1.26,1.55) | 1.44(1.32,1.57) | 2.45(1.55,5.17) |
| 95 | 1.48(1.05,2.39) | 1.07(0.96,1.2) | 1.11(1.01,1.22) | 1.89(1.13,4.84) |
| 96 | 1.04(0.71,1.82) | 0.75(0.67,0.86) | 0.79(0.72,0.88) | 1.35(0.74,4.58) |
| 97 | 0.66(0.43,1.29) | 0.48(0.43,0.56) | 0.51(0.46,0.57) | 0.87(0.43,4.15) |
| 98 | 0.35(0.22,0.79) | 0.26(0.23,0.3) | 0.28(0.25,0.32) | 0.47(0.16,3.17) |
| 99 | 0.13(0.08,0.35) | 0.09(0.08,0.11) | 0.1(0.09,0.12) | 0.18(-1.02,1.44) |
| 100 | 0(0,0) | 0(0,0) | 0(0,0) | 0(0,0) |

The model was adjusted for sex and race.

Supplementary Table S10 Redistribution of types of AF under the ICD classification into new clusters.

|  | Cluster 1 | Cluster 2 | Cluster 3 | Cluster 4 | Cluster 5 |
| --- | --- | --- | --- | --- | --- |
| Paroxysmal atrial fibrillation | 44 (10.5%) | 81 (19.2%) | 152 (36.1%) | 94 (22.3%) | 50 (11.9%) |
| Persistent atrial fibrillation | 8 (13.8%) | 10 (17.2%) | 23 (39.7%) | 7 (12.1%) | 10 (17.2%) |
| Chronic atrial fibrillation | 4 (8.3%) | 12 (25%) | 15 (31.3%) | 7 (14.6%) | 10 (20.8%) |
| Unspecified atrial fibrillation | 530 (7.7%) | 1487 (21.7%) | 2715 (39.6%) | 1143 (16.7%) | 976 (14.2%) |

Supplementary Table S11. Hazard ratios of death and major complications for the five AF cluster, comparing with non-AF.

|  | Cluster 1 | Cluster 2 | Cluster 3 | Cluster 4 | Cluster 5 |
| --- | --- | --- | --- | --- | --- |
| All-cause death | 1.41(1.09,1.81) | 2.61(2.33,2.94) | 2.73(2.51,2.97) | 2.74(2.42,3.11) | 4.55(4.06,5.09) |
| Cardiovascular death | 1.98(1.25,3.15) | 5.07(4.2,6.11) | 5.37(4.72,6.12) | 5.44(4.47,6.62) | 7.71(6.36,9.34) |
| AMI | 2.05(1.61,2.61) | 3.76(3.34,4.23) | 4.1(3.77,4.44) | 2.98(2.59,3.43) | 4.68(4.11,5.34) |
| HF | 7.43(6.28,8.79) | 9.26(8.4,10.21) | 9.33(8.69,10.02) | 9.06(8.13,10.1) | 13.37(12.05,14.83) |
| CIS | 2.96(2.12,4.15) | 4.85(4.1,5.74) | 4.52(3.98,5.13) | 4.3(3.53,5.23) | 6.22(5.15,7.51) |

Cox proportional hazards models were adjusted for sex, race. AMI, acute myocardial infarction, HF, heart failure, CIS, cerebral ischemic stroke.

Supplementary Table S12. Reductions of life expectency for participants in the five AF clusters comparing with non-AF participants.

| Age | Cluster 1 | Cluster 2 | Cluster 3 | Cluster 4 | Cluster 5 |
| --- | --- | --- | --- | --- | --- |
| 40 | 10.91(9.95,11.88) | 28.37(26.93,29.83) | 10.16(9.19,11.16) | 6.84(6.01,7.68) | 30.82(29.32,32.31) |
| 41 | 11.12(10.18,12.07) | 27.44(25.97,28.91) | 10.23(9.28,11.2) | 7.1(6.29,7.91) | 29.9(28.37,31.42) |
| 42 | 11.27(10.36,12.19) | 26.54(25.06,28.03) | 10.26(9.34,11.2) | 7.29(6.51,8.08) | 29.01(27.46,30.56) |
| 43 | 11.37(10.48,12.27) | 25.73(24.25,27.24) | 10.25(9.35,11.17) | 7.43(6.67,8.2) | 28.21(26.65,29.78) |
| 44 | 11.44(10.57,12.32) | 24.95(23.47,26.46) | 10.22(9.35,11.12) | 7.54(6.8,8.28) | 27.43(25.85,29.01) |
| 45 | 11.47(10.61,12.34) | 24.39(22.91,25.89) | 10.18(9.33,11.06) | 7.59(6.87,8.32) | 26.86(25.29,28.45) |
| 46 | 11.48(10.64,12.33) | 23.96(22.49,25.46) | 10.14(9.31,11.01) | 7.62(6.92,8.34) | 26.43(24.86,28.02) |
| 47 | 11.48(10.65,12.32) | 23.49(22.03,24.99) | 10.09(9.27,10.93) | 7.65(6.96,8.35) | 25.96(24.39,27.55) |
| 48 | 11.46(10.65,12.29) | 23.06(21.61,24.56) | 10.03(9.23,10.86) | 7.66(6.99,8.35) | 25.52(23.95,27.11) |
| 49 | 11.43(10.64,12.24) | 22.33(20.88,23.83) | 9.92(9.13,10.72) | 7.66(7.01,8.33) | 24.78(23.2,26.38) |
| 50 | 11.32(10.55,12.11) | 20.87(19.41,22.38) | 9.67(8.91,10.45) | 7.64(7.01,8.28) | 23.27(21.67,24.91) |
| 51 | 11.23(10.48,12) | 19.89(18.43,21.4) | 9.49(8.75,10.25) | 7.6(6.99,8.23) | 22.25(20.64,23.91) |
| 52 | 11.19(10.45,11.95) | 19.46(18.02,20.96) | 9.4(8.68,10.15) | 7.58(6.98,8.2) | 21.81(20.21,23.46) |
| 53 | 11.1(10.38,11.85) | 18.78(17.36,20.28) | 9.26(8.56,9.99) | 7.54(6.95,8.14) | 21.11(19.51,22.76) |
| 54 | 11.05(10.34,11.78) | 18.41(17,19.9) | 9.17(8.49,9.89) | 7.51(6.94,8.1) | 20.72(19.13,22.37) |
| 55 | 10.96(10.26,11.67) | 17.82(16.42,19.3) | 9.03(8.36,9.73) | 7.45(6.89,8.03) | 20.1(18.52,21.74) |
| 56 | 10.88(10.2,11.58) | 17.38(16,18.85) | 8.92(8.27,9.61) | 7.41(6.86,7.97) | 19.64(18.07,21.28) |
| 57 | 10.77(10.1,11.46) | 16.82(15.45,18.27) | 8.77(8.13,9.44) | 7.34(6.81,7.89) | 19.04(17.48,20.68) |
| 58 | 10.71(10.05,11.39) | 16.51(15.16,17.95) | 8.69(8.06,9.34) | 7.3(6.78,7.84) | 18.72(17.17,20.35) |
| 59 | 10.64(9.99,11.31) | 16.21(14.87,17.64) | 8.6(7.99,9.25) | 7.25(6.74,7.78) | 18.4(16.87,20.02) |
| 60 | 10.51(9.87,11.17) | 15.66(14.34,17.08) | 8.44(7.84,9.07) | 7.17(6.67,7.68) | 17.82(16.29,19.43) |
| 61 | 10.42(9.79,11.07) | 15.29(13.98,16.69) | 8.32(7.74,8.94) | 7.1(6.61,7.61) | 17.41(15.9,19.02) |
| 62 | 10.33(9.71,10.96) | 14.95(13.65,16.34) | 8.21(7.64,8.81) | 7.04(6.56,7.54) | 17.05(15.55,18.65) |
| 63 | 10.22(9.62,10.85) | 14.57(13.3,15.95) | 8.09(7.53,8.68) | 6.97(6.5,7.45) | 16.65(15.16,18.24) |
| 64 | 10.11(9.51,10.72) | 14.19(12.93,15.56) | 7.96(7.41,8.53) | 6.89(6.43,7.36) | 16.24(14.76,17.83) |
| 65 | 9.96(9.38,10.57) | 13.74(12.5,15.09) | 7.79(7.26,8.36) | 6.79(6.34,7.25) | 15.74(14.28,17.33) |
| 66 | 9.85(9.28,10.45) | 13.41(12.19,14.75) | 7.67(7.15,8.22) | 6.71(6.28,7.16) | 15.39(13.94,16.97) |
| 67 | 9.74(9.18,10.32) | 13.09(11.88,14.42) | 7.55(7.04,8.09) | 6.63(6.21,7.08) | 15.04(13.6,16.62) |
| 68 | 9.62(9.07,10.19) | 12.76(11.57,14.07) | 7.42(6.92,7.95) | 6.55(6.13,6.98) | 14.69(13.26,16.25) |
| 69 | 9.49(8.95,10.06) | 12.44(11.26,13.74) | 7.29(6.81,7.81) | 6.46(6.05,6.88) | 14.33(12.92,15.89) |
| 70 | 9.35(8.82,9.91) | 12.08(10.93,13.37) | 7.15(6.67,7.65) | 6.36(5.96,6.77) | 13.95(12.54,15.5) |
| 71 | 9.2(8.68,9.74) | 11.73(10.59,13) | 6.99(6.53,7.48) | 6.25(5.87,6.65) | 13.56(12.17,15.1) |
| 72 | 9.04(8.54,9.58) | 11.37(10.26,12.64) | 6.84(6.39,7.32) | 6.14(5.77,6.53) | 13.17(11.8,14.72) |
| 73 | 8.88(8.38,9.4) | 11.02(9.92,12.27) | 6.68(6.24,7.14) | 6.03(5.66,6.41) | 12.78(11.41,14.32) |
| 74 | 8.71(8.22,9.23) | 10.67(9.59,11.91) | 6.52(6.09,6.97) | 5.91(5.55,6.28) | 12.4(11.04,13.94) |
| 75 | 8.52(8.05,9.03) | 10.3(9.23,11.52) | 6.34(5.93,6.78) | 5.77(5.43,6.13) | 11.99(10.65,13.53) |
| 76 | 8.32(7.85,8.82) | 9.91(8.86,11.12) | 6.15(5.75,6.58) | 5.63(5.29,5.98) | 11.56(10.23,13.1) |
| 77 | 8.11(7.65,8.59) | 9.52(8.49,10.71) | 5.96(5.57,6.37) | 5.48(5.15,5.82) | 11.13(9.81,12.67) |
| 78 | 7.88(7.43,8.36) | 9.12(8.11,10.3) | 5.75(5.37,6.15) | 5.32(5,5.64) | 10.69(9.38,12.23) |
| 79 | 7.63(7.2,8.1) | 8.7(7.71,9.87) | 5.53(5.17,5.92) | 5.14(4.84,5.46) | 10.22(8.93,11.77) |
| 80 | 7.37(6.94,7.82) | 8.27(7.3,9.42) | 5.3(4.95,5.67) | 4.95(4.66,5.26) | 9.74(8.46,11.28) |
| 81 | 7.08(6.67,7.53) | 7.83(6.88,8.96) | 5.06(4.72,5.42) | 4.75(4.47,5.04) | 9.25(7.98,10.8) |
| 82 | 6.79(6.39,7.23) | 7.39(6.46,8.51) | 4.81(4.48,5.16) | 4.55(4.28,4.83) | 8.75(7.51,10.31) |
| 83 | 6.48(6.09,6.9) | 6.93(6.03,8.04) | 4.55(4.24,4.88) | 4.32(4.07,4.59) | 8.24(7.01,9.81) |
| 84 | 6.16(5.78,6.57) | 6.48(5.6,7.56) | 4.28(3.98,4.6) | 4.09(3.85,4.35) | 7.73(6.52,9.3) |
| 85 | 5.81(5.44,6.21) | 6.01(5.16,7.07) | 4(3.72,4.3) | 3.85(3.62,4.09) | 7.2(6.01,8.78) |
| 86 | 5.45(5.09,5.84) | 5.53(4.72,6.57) | 3.71(3.45,3.99) | 3.59(3.37,3.82) | 6.66(5.49,8.24) |
| 87 | 5.06(4.72,5.44) | 5.04(4.27,6.06) | 3.41(3.16,3.67) | 3.32(3.12,3.54) | 6.1(4.96,7.69) |
| 88 | 4.65(4.32,5.02) | 4.54(3.8,5.52) | 3.09(2.86,3.34) | 3.03(2.84,3.24) | 5.52(4.42,7.12) |
| 89 | 4.23(3.91,4.58) | 4.04(3.35,4.99) | 2.77(2.56,3) | 2.74(2.56,2.93) | 4.95(3.89,6.55) |
| 90 | 3.79(3.49,4.13) | 3.54(2.9,4.45) | 2.45(2.25,2.66) | 2.44(2.28,2.61) | 4.36(3.36,5.97) |
| 91 | 3.36(3.08,3.68) | 3.07(2.48,3.93) | 2.13(1.96,2.33) | 2.14(2,2.3) | 3.81(2.87,5.41) |
| 92 | 2.91(2.65,3.21) | 2.59(2.06,3.4) | 1.81(1.66,1.98) | 1.83(1.7,1.98) | 3.24(2.37,4.82) |
| 93 | 2.46(2.22,2.74) | 2.13(1.66,2.87) | 1.5(1.37,1.65) | 1.53(1.42,1.66) | 2.69(1.91,4.25) |
| 94 | 1.99(1.78,2.25) | 1.68(1.28,2.35) | 1.19(1.08,1.32) | 1.23(1.13,1.33) | 2.14(1.47,3.66) |
| 95 | 1.55(1.37,1.78) | 1.27(0.95,1.84) | 0.91(0.82,1.01) | 0.94(0.87,1.03) | 1.64(1.07,3.09) |
| 96 | 1.12(0.98,1.31) | 0.89(0.64,1.35) | 0.64(0.57,0.71) | 0.67(0.61,0.73) | 1.16(0.72,2.5) |
| 97 | 0.74(0.63,0.88) | 0.56(0.4,0.9) | 0.41(0.36,0.46) | 0.43(0.39,0.48) | 0.74(0.44,1.93) |
| 98 | 0.41(0.34,0.5) | 0.3(0.2,0.51) | 0.22(0.19,0.25) | 0.23(0.21,0.26) | 0.4(0.22,1.32) |
| 99 | 0.15(0.12,0.19) | 0.11(0.07,0.2) | 0.08(0.07,0.09) | 0.09(0.08,0.1) | 0.15(0.07,0.64) |
| 100 | 0(0,0) | 0(0,0) | 0(0,0) | 0(0,0) | 0(0,0) |

The model was adjusted for sex and race.

Supplementary Table S13. Genetic associations of risk variants for AF with the five clusters.

| SNP | Nearest GENE | Cluster 1 | | Cluster 2 | | Cluster 3 | | Cluster 4 | | Cluster 5 | | P_interaction_ |
| --- | --- | --- | --- | --- | --- | --- | --- | --- | --- | --- | --- | --- |
|  |  | BETA | P | BETA | P | BETA | P | BETA | P | BETA | P |  |
| rs10165883 | SNRNP27 | -6.75E-06 | 0.92738 | 0.000204 | 0.09419 | 0.00021683 | 0.18573 | 0.0002389 | 0.0273 | 0.0001466 | 0.13863 | 0.00205 |
| rs10213171 | ARHGAP10 | -6.35E-05 | 0.87218 | -0.000355 | 0.58329 | 0.00177943 | 0.04024 | -0.000106 | 0.85375 | 0.0004157 | 0.4285 | 0.056148 |
| rs1044258 | C10orf76 | -0.000129 | 0.12253 | 8.72E-05 | 0.52641 | 0.0003811 | 0.03898 | 0.0005677 | 3.24E-06 | 0.000341 | 0.00224 | 3.33E-07 |
| rs10520260 | HAND2 | -8.46E-05 | 0.36536 | 0.0002579 | 0.09359 | 0.00017435 | 0.39843 | 0.0003581 | 0.00798 | 0.0002281 | 0.06622 | 0.00049 |
| rs10753933 | PPFIA4 | 3.50E-05 | 0.63289 | 0.0003455 | 0.00411 | 0.00037856 | 0.01906 | 0.0002521 | 0.01834 | 0.000386 | 7.86E-05 | 5.65E-08 |
| rs10760361 | PSMB7 | 2.49E-05 | 0.75718 | -4.50E-06 | 0.97283 | 0.00058661 | 0.00093 | 4.90E-05 | 0.6753 | -7.44E-05 | 0.48734 | 0.003546 |
| rs10842383 | LINC00477 | 0.0003658 | 0.02968 | 0.0001136 | 0.68103 | 0.00052046 | 0.16059 | 0.0002805 | 0.25202 | 0.0001582 | 0.48041 | 0.014639 |
| rs10873299 | LRRC74 | 8.82E-06 | 0.91191 | 0.0002637 | 0.04362 | 0.00058189 | 0.0009 | 0.0002146 | 0.06425 | 0.0002617 | 0.01373 | 3.08E-06 |
| rs11001667 | C10orf11 | 7.49E-05 | 0.61695 | 1.12E-05 | 0.96355 | 0.00061695 | 0.06135 | 6.51E-05 | 0.76492 | 0.0001427 | 0.47339 | 0.040874 |
| rs11180703 | KRR1 | 4.92E-05 | 0.50531 | 9.01E-05 | 0.45798 | 0.00032923 | 0.04378 | 0.0001182 | 0.27325 | 0.0002471 | 0.01235 | 0.001419 |
| rs11264280 | KCNN3 | 0.0001797 | 0.04474 | 0.0007819 | 8.81E-08 | 0.00146185 | 8.80E-14 | 0.0005184 | 6.43E-05 | 0.0004884 | 4.03E-05 | 5.57E-26 |
| rs113819537 | SSPN | 0.0001703 | 0.11307 | 0.0005047 | 0.00427 | 0.00070228 | 0.00305 | 0.0001378 | 0.37965 | 0.0002076 | 0.14732 | 7.92E-06 |
| rs11598047 | NEURL | 0.000221 | 0.19152 | 0.0011186 | 5.67E-05 | 0.00073128 | 0.05006 | 0.0006248 | 0.01127 | 0.0004801 | 0.03322 | 2.65E-07 |
| rs11768850 | SUN1 | 3.36E-05 | 0.65198 | 0.0002365 | 0.05313 | 8.72E-05 | 0.59503 | 3.86E-05 | 0.72233 | 6.56E-05 | 0.50805 | 0.060317 |
| rs11773845 | CAV1 | 0.0002986 | 7.40E-05 | 0.0005669 | 4.58E-06 | 0.0008628 | 2.05E-07 | 0.00012 | 0.2744 | 0.0001793 | 0.07394 | 9.56E-15 |
| rs117984853 | UST | 0.0002157 | 0.45628 | 0.0011806 | 0.01291 | 0.00136037 | 0.0328 | -0.000502 | 0.23457 | 0.0006379 | 0.0979 | 0.000798 |
| rs12044963 | KCND3 | 0.0001367 | 0.60755 | 0.0005981 | 0.17095 | 0.00068191 | 0.24496 | 0.0007747 | 0.04567 | 0.0002744 | 0.4389 | 0.008321 |
| rs12208899 | EYA4 | 0.0001324 | 0.27764 | 0.0005148 | 0.01003 | 0.00061187 | 0.02266 | 0.0002919 | 0.09992 | 8.25E-05 | 0.61169 | 0.000188 |
| rs12298484 | DNAH10 | 0.0002144 | 0.01209 | 0.0002742 | 0.05078 | 0.00041142 | 0.02908 | 0.0002197 | 0.07777 | -2.23E-05 | 0.84493 | 0.000368 |
| rs12591736 | TLE3 | -0.00015 | 0.41807 | 0.0002264 | 0.45535 | -0.0001149 | 0.7785 | 0.0003226 | 0.23259 | 0.0001438 | 0.5597 | 0.226645 |
| rs12809354 | PKP2 | 0.0001734 | 0.33905 | 0.0004975 | 0.09512 | 0.00125616 | 0.00168 | 0.0005803 | 0.02811 | 0.0004343 | 0.07235 | 8.98E-06 |
| rs12810346 | TBX3 | 0.0002266 | 0.24107 | -0.000366 | 0.25072 | 0.00029288 | 0.4941 | -4.31E-05 | 0.87922 | 0.000653 | 0.01135 | 0.045599 |
| rs12908004 | ARNT2 | 0.0002204 | 0.16615 | 0.000408 | 0.11835 | 7.66E-06 | 0.98257 | 0.0001412 | 0.54277 | 0.0005402 | 0.01079 | 0.001609 |
| rs12908437 | IGF1R | -1.32E-05 | 0.86856 | 6.10E-05 | 0.64181 | 4.06E-05 | 0.81816 | -6.91E-05 | 0.55321 | 0.0001295 | 0.22412 | 0.331695 |
| rs12992412 | MBD5 | 8.69E-05 | 0.28382 | 0.0001265 | 0.34282 | 0.00055437 | 0.00195 | 0.000214 | 0.0704 | 0.0001934 | 0.07419 | 4.24E-05 |
| rs1307274 | NUDT3 | 0.0008366 | 0.01839 | 0.0007607 | 0.19264 | 0.0006488 | 0.40775 | 0.0007667 | 0.13833 | 0.0011602 | 0.01413 | 9.54E-05 |
| rs13191450 | GJA1 | 0.000118 | 0.14932 | 0.0004019 | 0.00278 | 0.00042243 | 0.01911 | 0.0001659 | 0.16391 | 0.0001679 | 0.12317 | 4.20E-05 |
| rs146518726 | C1orf185 | -0.000582 | 0.64336 | -0.001632 | 0.43012 | -0.0029325 | 0.29179 | -0.001226 | 0.50324 | 0.0040486 | 0.0152 | 0.487803 |
| rs17079881 | SLC35F1 | -6.08E-06 | 0.97489 | 0.0002391 | 0.45191 | 0.00178039 | 2.99E-05 | 0.0006573 | 0.01973 | 0.0001147 | 0.65688 | 3.04E-06 |
| rs174048 | NR3C1 | 5.64E-05 | 0.72574 | -8.38E-05 | 0.75082 | 0.00065212 | 0.06559 | 0.000317 | 0.17587 | 0.0001333 | 0.53355 | 0.022026 |
| rs17490701 | PHLDB2 | 0.0002369 | 0.17919 | 1.24E-05 | 0.9658 | 0.00051912 | 0.18055 | 0.0001304 | 0.61109 | -8.35E-05 | 0.72068 | 0.09093 |
| rs1822273 | NAV2 | 0.000209 | 0.06292 | 0.0003311 | 0.07304 | 0.00099332 | 6.17E-05 | 1.91E-05 | 0.90723 | 0.0002026 | 0.17663 | 1.61E-06 |
| rs187585530 | UBE4B | -0.000506 | 0.94356 | -0.001981 | 0.8663 | -0.0042467 | 0.78785 | -0.00169 | 0.87135 | -0.001523 | 0.87308 | 0.746444 |
| rs210632 | GOPC | 3.27E-05 | 0.76213 | 9.86E-05 | 0.57741 | 0.00042551 | 0.07332 | 0.0001596 | 0.31016 | 5.98E-05 | 0.67695 | 0.031069 |
| rs2129977 | PITX2 | 0.0010089 | 4.89E-15 | 0.0020606 | 1.85E-22 | 0.00356029 | 2.56E-36 | 0.0018736 | 1.58E-23 | 0.0015124 | 1.13E-18 | 1.52E-86 |
| rs2145274 | CASC20 | -0.000191 | 0.54594 | 0.0004347 | 0.40236 | 0.00110902 | 0.11342 | 0.0004897 | 0.28796 | 0.0002856 | 0.49933 | 0.036029 |
| rs2145587 | AKAP6 | -5.99E-05 | 0.57341 | 0.000317 | 0.07017 | 0.00058849 | 0.01229 | 0.0003161 | 0.04176 | 0.0004446 | 0.00172 | 2.08E-05 |
| rs2286466 | RPS2 | 8.07E-06 | 0.95254 | 0.0001318 | 0.55382 | 0.00012284 | 0.68125 | 0.0004523 | 0.0222 | 7.13E-05 | 0.69333 | 0.047208 |
| rs2306272 | LRIG1 | 0.0001216 | 0.20045 | 0.0006068 | 9.87E-05 | -1.47E-05 | 0.94414 | 0.0001555 | 0.26105 | 0.0001415 | 0.26308 | 5.64E-05 |
| rs2359171 | ZFHX3 | 0.0003318 | 0.02791 | 0.0011624 | 2.71E-06 | 0.00173702 | 1.73E-07 | 0.000418 | 0.05754 | 0.0011117 | 3.16E-08 | 2.81E-17 |
| rs242557 | MAPT | 0.0001812 | 0.02045 | -2.93E-05 | 0.81939 | 0.00045475 | 0.00831 | 6.26E-05 | 0.58256 | 3.85E-05 | 0.71158 | 0.001377 |
| rs2540949 | CEP68 | 4.02E-05 | 0.60949 | 0.0001036 | 0.42233 | 0.00026147 | 0.13165 | 0.0002979 | 0.0094 | 0.0001575 | 0.1333 | 0.001151 |
| rs2738413 | SYNE2 | 4.59E-05 | 0.52618 | 0.0003907 | 0.00102 | 0.0003013 | 0.05932 | 0.0002864 | 0.00668 | 0.0001651 | 0.08751 | 4.94E-06 |
| rs2834618 | LOC100506385 | 0.0002767 | 0.29789 | 0.0001316 | 0.76292 | 0.00091413 | 0.11933 | -0.000482 | 0.21347 | 0.0001169 | 0.7415 | 0.093595 |
| rs28631169 | MYH7 | 9.58E-05 | 0.45971 | 0.0003318 | 0.11876 | 0.00126209 | 9.75E-06 | 0.0002883 | 0.12679 | 8.14E-05 | 0.63702 | 9.92E-07 |
| rs295114 | SPATS2L | 5.93E-05 | 0.42802 | 0.0001214 | 0.32409 | 0.0003217 | 0.0513 | 0.0001665 | 0.12763 | 0.0002856 | 0.00419 | 0.00048 |
| rs3176326 | CDKN1A | 0.0001405 | 0.29055 | -0.00022 | 0.3123 | 0.00080272 | 0.00604 | 1.52E-05 | 0.93751 | -2.37E-05 | 0.89349 | 0.011129 |
| rs34750263 | WNT8A | 0.0001038 | 0.24612 | 0.000378 | 0.01012 | 0.00079627 | 5.39E-05 | 0.000279 | 0.03234 | 0.000357 | 0.00276 | 7.08E-09 |
| rs34969716 | KDM1B | 3.64E-05 | 0.7214 | 0.0002062 | 0.21354 | 0.00033983 | 0.12688 | 0.0002523 | 0.08518 | 0.0002878 | 0.03319 | 0.00318 |
| rs35006907 | MTSS1 | 0.0002127 | 0.01846 | 0.0001235 | 0.40389 | 0.00025949 | 0.19148 | -7.53E-05 | 0.56683 | 0.0001182 | 0.32521 | 0.02362 |
| rs35349325 | BEST3 | 2.74E-05 | 0.70551 | 0.0001674 | 0.15986 | 0.00016209 | 0.31018 | 5.07E-05 | 0.63096 | 0.0001914 | 0.04782 | 0.009924 |
| rs35504893 | TTN | 0.0001423 | 0.29477 | 0.0001846 | 0.40759 | 0.00045731 | 0.12648 | 0.0005316 | 0.00719 | 0.0002357 | 0.19244 | 0.000952 |
| rs3822259 | WDR1 | -1.09E-05 | 0.90267 | -2.40E-05 | 0.86985 | 0.00055662 | 0.00459 | -5.48E-05 | 0.67142 | 2.94E-05 | 0.80371 | 0.017703 |
| rs3960788 | SLC9B1 | 0.0001369 | 0.06864 | 0.0002058 | 0.09511 | 0.00030963 | 0.06156 | 5.69E-05 | 0.60352 | -9.53E-05 | 0.34098 | 0.006357 |
| rs4385527 | C9orf3 | 7.91E-05 | 0.30495 | 0.0002338 | 0.06408 | 0.00067141 | 7.31E-05 | 0.0003425 | 0.00224 | 0.0001623 | 0.1128 | 4.14E-08 |
| rs4484922 | CASQ2 | 1.50E-05 | 0.87137 | -0.000129 | 0.39636 | 0.00028713 | 0.15833 | -7.53E-06 | 0.95534 | -1.21E-05 | 0.92153 | 0.263743 |
| rs465276 | TUBA8 | 2.01E-06 | 0.97961 | 2.56E-05 | 0.84374 | 0.00048835 | 0.00511 | 7.00E-05 | 0.54389 | 0.0001355 | 0.19898 | 0.002505 |
| rs4743034 | ZNF462 | 0.0002474 | 0.03075 | 2.53E-05 | 0.89297 | 0.00025824 | 0.3063 | 6.82E-05 | 0.68297 | 0.0001423 | 0.35088 | 0.052612 |
| rs4855075 | GNB4 | 0.0004895 | 0.00789 | -2.25E-05 | 0.94094 | 0.00045666 | 0.26151 | -4.79E-05 | 0.8586 | 0.0003303 | 0.17957 | 0.014867 |
| rs4951261 | NUCKS1 | 3.19E-05 | 0.67945 | -0.00017 | 0.18145 | 0.00040852 | 0.01642 | 0.000252 | 0.02517 | 0.0001277 | 0.2144 | 0.0014 |
| rs4977397 | SLC24A2 | 1.62E-05 | 0.83225 | 1.45E-05 | 0.90815 | 0.0002385 | 0.15866 | 0.0001353 | 0.22656 | 9.99E-05 | 0.32863 | 0.044341 |
| rs55734480 | DGKB | -4.39E-06 | 0.96568 | 0.0001622 | 0.33198 | 0.00030658 | 0.17237 | 0.0001958 | 0.1877 | 0.0001613 | 0.2349 | 0.027801 |
| rs55754224 | CAMK2D | -0.000111 | 0.298 | 0.0002028 | 0.24569 | 0.00018175 | 0.4384 | 9.87E-05 | 0.5239 | 3.75E-05 | 0.79109 | 0.151989 |
| rs55985730 | OPN1SW | -0.0006 | 0.22006 | 0.0014339 | 0.07454 | 0.00045547 | 0.6729 | 0.0006224 | 0.38289 | 0.000459 | 0.482 | 0.050303 |
| rs60212594 | WIPF1 | 0.0001345 | 0.44594 | 0.0001388 | 0.63198 | 0.00043426 | 0.26466 | 3.35E-06 | 0.98962 | 0.0002493 | 0.28893 | 0.105035 |
| rs608930 | GORAB | 0.0001218 | 0.10144 | 0.0005861 | 1.55E-06 | 0.00048772 | 0.00289 | 0.0001982 | 0.0671 | 0.0003529 | 0.00036 | 4.59E-11 |
| rs62011291 | USP3 | 0.0001586 | 0.19852 | 0.0003061 | 0.12985 | 0.00053941 | 0.04681 | 0.0003746 | 0.03667 | 0.0002486 | 0.13022 | 0.000748 |
| rs62483627 | COG5 | -0.000122 | 0.29091 | 0.0004628 | 0.01505 | 0.00032893 | 0.1983 | 0.0003203 | 0.05733 | 3.15E-05 | 0.83848 | 0.00325 |
| rs62521286 | FBX032 | 0.0002534 | 0.49128 | 0.0001436 | 0.81227 | 0.00075892 | 0.34948 | 0.0007242 | 0.17688 | 4.53E-05 | 0.92643 | 0.117729 |
| rs6462078 | CREB5 | -8.40E-05 | 0.41993 | 0.0001741 | 0.30945 | 0.00013802 | 0.54822 | -3.52E-05 | 0.8162 | -9.11E-05 | 0.51187 | 0.432834 |
| rs6546620 | KIF3C | 1.54E-05 | 0.91738 | 0.0003838 | 0.11496 | 0.00085373 | 0.00964 | 0.0002143 | 0.32314 | 7.36E-05 | 0.71259 | 0.001444 |
| rs6742276 | XP01 | 9.78E-05 | 0.20257 | 5.40E-05 | 0.66783 | 0.00036229 | 0.03213 | 0.000177 | 0.11325 | 9.60E-05 | 0.34756 | 0.002691 |
| rs6790396 | SCN10A | 0.0001454 | 0.05533 | 0.0001444 | 0.24666 | 0.00025769 | 0.12361 | 0.0002941 | 0.00782 | 0.0002169 | 0.032 | 0.00017 |
| rs6810325 | CAND2 | 4.82E-05 | 0.56755 | 4.84E-05 | 0.7266 | 0.00049817 | 0.00737 | 0.0002243 | 0.06799 | 0.0002369 | 0.03497 | 0.000198 |
| rs6882776 | NKX2–5 | 0.0001332 | 0.17123 | 0.0004602 | 0.00384 | 0.00054889 | 0.01041 | 0.0001769 | 0.21061 | 4.11E-05 | 0.75018 | 5.94E-05 |
| rs6907805 | CGA | 8.17E-05 | 0.25622 | 0.0001689 | 0.1526 | 0.0001487 | 0.34828 | 0.0001681 | 0.10897 | 0.000142 | 0.13802 | 0.00993 |
| rs6993266 | PTK2 | 4.98E-05 | 0.49384 | 0.0002648 | 0.02656 | 0.0003503 | 0.02921 | 5.44E-05 | 0.60861 | 0.0001829 | 0.05928 | 0.000532 |
| rs716845 | KCNN2 | -8.31E-05 | 0.4012 | 0.0001586 | 0.32831 | 0.00022083 | 0.31042 | 0.0003168 | 0.02788 | 3.86E-06 | 0.97663 | 0.024779 |
| rs7219869 | KCNJ2 | -2.88E-07 | 0.9969 | 0.0001359 | 0.26694 | 5.31E-05 | 0.7463 | 8.13E-05 | 0.45212 | 7.39E-05 | 0.45682 | 0.129612 |
| rs7269123 | C20orf166 | 0.0001226 | 0.11006 | 0.0001317 | 0.2924 | 0.00014796 | 0.3806 | -1.59E-05 | 0.88647 | 0.0001388 | 0.17193 | 0.064984 |
| rs72700114 | METTL11B | -0.000171 | 0.64682 | 0.0028812 | 2.28E-06 | 0.00223742 | 0.00633 | 0.0025223 | 3.42E-06 | 0.0011937 | 0.01594 | 8.33E-13 |
| rs72811294 | MYOCD | 0.000225 | 0.32027 | 0.0006671 | 0.07282 | 0.00077294 | 0.12156 | 0.0005408 | 0.10075 | -0.000519 | 0.08483 | 0.010028 |
| rs72926475 | REEP1 | -1.94E-05 | 0.92568 | 0.0004021 | 0.23931 | -0.0004931 | 0.28256 | -3.73E-05 | 0.90197 | -7.55E-05 | 0.78568 | 0.584351 |
| rs73032363 | THRB | 0.0002104 | 0.02577 | 0.0003531 | 0.02306 | 0.00029846 | 0.15358 | 0.0001519 | 0.27136 | -9.54E-05 | 0.44992 | 0.001857 |
| rs73241997 | SNX6,CFL2 | -4.74E-05 | 0.77023 | 0.0004932 | 0.0637 | 0.00046205 | 0.19588 | -0.000216 | 0.36159 | -0.000228 | 0.29166 | 0.091476 |
| rs73366713 | ATXN1 | 0.0002657 | 0.14269 | -3.70E-05 | 0.9013 | 0.00081989 | 0.04038 | 0.000333 | 0.20712 | 0.0005204 | 0.03154 | 0.001535 |
| rs74022964 | HCN4 | 1.02E-05 | 0.95072 | 0.0005722 | 0.03566 | 0.00074123 | 0.04246 | 0.000186 | 0.44126 | 7.75E-05 | 0.72633 | 0.003873 |
| rs7460121 | MIR30B | 0.0002172 | 0.38994 | 7.31E-06 | 0.98595 | 0.00160195 | 0.00397 | 3.43E-05 | 0.92569 | 0.0003763 | 0.26328 | 0.001907 |
| rs74910854 | GTF2I | -0.000145 | 0.69987 | 0.0006299 | 0.30687 | 0.00289403 | 0.00045 | -0.000375 | 0.49257 | -0.000624 | 0.21242 | 0.002771 |
| rs7508 | ASAH1 | 0.0002036 | 0.04051 | 0.0001089 | 0.50371 | 0.00033851 | 0.12189 | 0.0002336 | 0.10512 | 0.0001702 | 0.19823 | 0.005231 |
| rs76097649 | KCNJ5 | 0.0008611 | 0.00267 | 0.0016836 | 0.00034 | 0.00045068 | 0.47587 | 0.0010921 | 0.00892 | 0.0006565 | 0.08559 | 9.02E-08 |
| rs7789146 | KCNH2 | 6.57E-05 | 0.64569 | 4.44E-05 | 0.84978 | -0.000106 | 0.73652 | 0.0003237 | 0.12023 | 0.0002553 | 0.17998 | 0.055798 |
| rs7846485 | XP07 | 0.0002601 | 0.19601 | 0.0006205 | 0.06005 | -0.0008417 | 0.05723 | -0.000184 | 0.53039 | 9.13E-06 | 0.97281 | 0.159322 |
| rs79187193 | GJA5 | 7.77E-06 | 0.99019 | -0.000248 | 0.81065 | 0.0005319 | 0.70256 | 6.37E-05 | 0.94482 | 0.0011203 | 0.18255 | 0.322647 |
| rs7919685 | REEP3 | 7.57E-06 | 0.91619 | 3.31E-05 | 0.77947 | 0.00058793 | 0.00021 | 0.0001426 | 0.17423 | 0.00026 | 0.00673 | 4.37E-06 |
| rs7978685 | NACA | 5.39E-05 | 0.58766 | 7.40E-05 | 0.65076 | 0.00033255 | 0.12952 | 0.0005187 | 0.00034 | 0.0002097 | 0.11317 | 0.000177 |
| rs8073937 | POLR2A | 0.0003024 | 0.00027 | 5.28E-05 | 0.69807 | 0.00027539 | 0.13207 | 0.0002337 | 0.05306 | 8.39E-05 | 0.4477 | 0.000229 |
| rs880315 | CASZ1 | 3.30E-05 | 0.69979 | -6.19E-05 | 0.6613 | 0.00017844 | 0.34728 | 0.0001381 | 0.27123 | -8.31E-05 | 0.46646 | 0.241117 |
| rs883079 | TBX5 | 0.0001494 | 0.12894 | 0.0004834 | 0.00278 | 0.00081161 | 0.00018 | 0.0004828 | 0.00076 | 0.000436 | 0.00088 | 8.90E-10 |
| rs949078 | sorli | 8.93E-05 | 0.39703 | 0.0002369 | 0.17163 | 0.00046453 | 0.04575 | -4.91E-05 | 0.7493 | 6.07E-06 | 0.96552 | 0.014161 |
| rs9580438 | BASP1P1 | 0.0001224 | 0.1551 | 0.0001193 | 0.39693 | 5.97E-06 | 0.97482 | 0.0001019 | 0.41438 | 0.0002475 | 0.02979 | 0.007137 |
| rs9872035 | PAK2 | 4.89E-05 | 0.50674 | 0.0002975 | 0.01379 | 8.14E-05 | 0.61559 | 0.0001039 | 0.33331 | 0.0001204 | 0.21983 | 0.003909 |
| rs9953366 | SMAD7 | -9.15E-05 | 0.3201 | 0.000116 | 0.43976 | 6.39E-05 | 0.75173 | 0.000191 | 0.15536 | 9.04E-05 | 0.46277 | 0.103284 |

The generalized linear model was used to estimate the associations of each risk variant for AF with the five clusters, after adjustment of age, sex, assessment centres, top 10 principal components of ancestry, and dominance-deviation. The P_interaction_ was based on the assessment of the significance in the differences of associations between the specific risk variant and the five clusters.

Supplementary Table S14. The difference in participant number between origin cluster and re-cluster

|  |  | Re-cluster | | | | |
| --- | --- | --- | --- | --- | --- | --- |
| Cluster | Participants | Cluster1 | Cluster2 | Cluster3 | Cluster4 | Cluster5 |
| Cluster1 | 587 | 551 | 0 | 5 | 0 | 31 |
| Cluster2 | 1593 | 114 | 1332 | 2 | 111 | 34 |
| Cluster3 | 2912 | 324 | 11 | 2475 | 52 | 50 |
| Cluster4 | 1253 | 58 | 65 | 0 | 1080 | 50 |
| Cluster5 | 1046 | 35 | 0 | 2 | 14 | 995 |

Supplementary Table S15. Hazard ratios of death and complications associated with AF cluster in participants with newly-onset AF in 2 years after baseline.

|  | HR (95%CI) for all-cause death | | | | |
| --- | --- | --- | --- | --- | --- |
| Reference | Cluster 1 | Cluster 2 | Cluster 3 | Cluster 4 | Cluster 5 |
| Cluster 1 | 1 | 1.69(1.21,2.35) | 1.43(1.04,1.95) | 1.16(0.82,1.63) | 2.43(1.76,3.34) |
| Cluster 2 | 0.59(0.43,0.83) | 1 | 0.85(0.66,1.08) | 0.69(0.52,0.91) | 1.44(1.12,1.85) |
| Cluster 3 | 0.7(0.51,0.96) | 1.18(0.92,1.51) | 1 | 0.81(0.63,1.05) | 1.7(1.35,2.14) |
| Cluster 4 | 0.86(0.61,1.22) | 1.46(1.1,1.93) | 1.23(0.95,1.6) | 1 | 2.1(1.61,2.74) |
| Cluster 5 | 0.41(0.3,0.57) | 0.69(0.54,0.89) | 0.59(0.47,0.74) | 0.48(0.37,0.62) | 1 |
|  | HR (95%CI) for cardiovascular death | | | | |
| Reference | Cluster 1 | Cluster 2 | Cluster 3 | Cluster 4 | Cluster 5 |
| Cluster 1 | 1 | 1.72(1.12,2.64) | 1.43(0.95,2.15) | 1.03(0.65,1.62) | 2.62(1.74,3.95) |
| Cluster 2 | 0.58(0.38,0.89) | 1 | 0.83(0.6,1.14) | 0.6(0.41,0.87) | 1.52(1.11,2.1) |
| Cluster 3 | 0.7(0.47,1.05) | 1.2(0.88,1.66) | 1 | 0.72(0.51,1.02) | 1.84(1.37,2.46) |
| Cluster 4 | 0.97(0.62,1.53) | 1.67(1.15,2.44) | 1.39(0.98,1.97) | 1 | 2.55(1.79,3.64) |
| Cluster 5 | 0.38(0.25,0.57) | 0.66(0.48,0.9) | 0.54(0.41,0.73) | 0.39(0.27,0.56) | 1 |

|  | HR (95%CI) for AMI | | | | |
| --- | --- | --- | --- | --- | --- |
| Reference | Cluster 1 | Cluster 2 | Cluster 3 | Cluster 4 | Cluster 5 |
| Cluster 1 | 1 | 1.09(0.46,2.56) | 1.26(0.59,2.71) | 1.19(0.52,2.69) | 2.25(1.04,4.9) |
| Cluster 2 | 0.92(0.39,2.16) | 1 | 1.16(0.59,2.29) | 1.09(0.52,2.28) | 2.07(1.04,4.12) |
| Cluster 3 | 0.79(0.37,1.71) | 0.86(0.44,1.71) | 1 | 0.94(0.5,1.77) | 1.79(1.01,3.19) |
| Cluster 4 | 0.84(0.37,1.91) | 0.92(0.44,1.92) | 1.06(0.56,2) | 1 | 1.90(1.01,3.62) |
| Cluster 5 | 0.44(0.2,0.97) | 0.48(0.24,0.96) | 0.56(0.31,0.99) | 0.53(0.28,0.99) | 1 |
|  | HR (95%CI) for HF | | | | |
| Reference | Cluster 1 | Cluster 2 | Cluster 3 | Cluster 4 | Cluster 5 |
| Cluster 1 | 1 | 2.27(1.48,3.46) | 2.02(1.35,3.03) | 1.95(1.28,2.98) | 3.14(2.07,4.77) |
| Cluster 2 | 0.44(0.29,0.67) | 1 | 0.89(0.67,1.18) | 0.86(0.63,1.17) | 1.39(1.03,1.86) |
| Cluster 3 | 0.49(0.33,0.74) | 1.12(0.85,1.48) | 1 | 0.97(0.73,1.28) | 1.56(1.19,2.04) |
| Cluster 4 | 0.51(0.34,0.78) | 1.16(0.86,1.58) | 1.04(0.78,1.37) | 1 | 1.61(1.2,2.17) |
| Cluster 5 | 0.32(0.21,0.48) | 0.72(0.54,0.97) | 0.64(0.49,0.84) | 0.62(0.46,0.83) | 1 |
|  | HR (95%CI) for CIS | | | | |
| Reference | Cluster 1 | Cluster 2 | Cluster 3 | Cluster 4 | Cluster 5 |
| Cluster 1 | 1 | 6.91(2.09,22.85) | 5.99(1.85,19.43) | 9.24(2.85,29.9) | 8.11(2.45,26.82) |
| Cluster 2 | 0.14(0.04,0.48) | 1 | 0.87(0.52,1.43) | 1.34(0.81,2.19) | 1.17(0.68,2.02) |
| Cluster 3 | 0.17(0.05,0.54) | 1.15(0.7,1.91) | 1 | 1.54(0.98,2.42) | 1.35(0.82,2.24) |
| Cluster 4 | 0.11(0.03,0.35) | 0.75(0.46,1.23) | 0.65(0.41,1.02) | 1 | 0.88(0.53,1.44) |
| Cluster 5 | 0.12(0.04,0.41) | 0.85(0.5,1.46) | 0.74(0.45,1.22) | 1.14(0.69,1.87) | 1 |

Cox proportional hazards models were adjusted for sex and race. AMI, acute myocardial infarction, HF, heart failure, CIS, cerebral ischemic stroke.

Supplementary Method 1. LASSO algorithm

The Cox LASSO model is a shrinkage technique that effectively selects a subset of variables from a large and potentially multicollinear set of predictors in regression analysis, leading to a more meaningful and interpretable set of predictors.

In the Cox LASSO regression model, we included 409,661 participants who were free of atrial fibrillation (AF) at baseline and provided complete data. Among them, 23,760 participants were diagnosed with AF during follow-up and coded as 1 for the outcome variable, while the remaining participants were coded as 0. To facilitate to merge variables in subsequent analyses, we set thresholds based on clinical significance and transformed continuous variables into binary categorical variables, as outlined in Supplemental Table 3. Following guidelines, we incorporated 25 variables associated with AF risk, comprising 18 lifestyle, metabolic factors, and comorbidities, along with 7 covariates (4 non-modifiable variables and 3 social factors).

LASSO achieves the optimal model by performing a continuous shrinking operation, minimizing regression coefficients to mitigate the risk of overfitting. As the value of lambda increases, the corresponding estimated parameters are increasingly shrunk, and when lambda reaches a certain threshold, some non-important variables will be compressed to zero, indicating their removal from the model. However, while reducing the number of variables, the model is subject to greater penalties, such as a decline in predictive capacity and goodness of fit. The C-index and the partial likelihood deviation can be selected for lasso regression of cox model ^1^. C-index is used to measure the discrimination of the model, which represents the predictive capacity of model. The partial likelihood deviation is used to calculate the mean squared error of the variables, which value is as small as possible to represent a good model fitting. The key characteristic of LASSO lies in its ability to induce sparsity by imposing a constraint that shrinks the sum of the absolute values of regression coefficients, thereby encouraging some coefficients to become exactly zero, effectively selecting the non-zero variables to remain in the model.

Supplementary Method 2. Factor analysis protocol

First, the variables were tested for applicability to exploratory factor analysis (EFA). All variables are considered to be independent of each other if the matrix of correlation coefficients formed by all the variables is a unit array (diagonal elements have a value of 1 while the rest of the elements have a value of 0). Bartlett's spherical test, prior to EFA, was adopt to test the difference between the matrix of correlation coefficients and a unit array to prove the existence of interactions between the variables. There was correlation between the 16 variables that were included (Chi-square = 4586, P < 0.001), indicating that the data were suitable for factor analysis. Subsequently, from the correlation matrix, a partial correlation matrix was calculated. The partial correlation coefficient is the independent correlation between two variables, controlling for the effects of other variables. Multicollinearity between variables is reflected by the partial correlation matrix. The KMO value is an indicator between 0 and 1, which measures the overall measure of correlation between all variables. A higher KMO value indicates a stronger correlation between the variables and is suitable for use in factor analysis. The KMO value of variables in our analysis was 0.67, which is considered acceptable (> 0.6).

Second, we used EFA with the algorithm of oblique (Geomin) rotation on 16 risk factors to establish a factor structure ^2^. We used the eigen value to determine the most appropriate number of factors to retain. In the process of factor analysis, the correlation or covariance matrix is calculated and decomposed into eigenvalues. The eigenvalues represent a measure of variance that reflects the number of factors present in the data and the ability of each factor to explain the variance of the data. For a data set with a sample size of M, the decomposition of all variables generates up to M eigenvalues. These eigenvalues are arranged in order from largest to smallest. The largest eigenvalue represents the first factor, which explains the largest variance in the original data. The second largest eigenvalue denotes the second principal component or second factor, which explains the second largest portion of the remaining variance, and so on. We chose the value corresponding to the horizontal coordinate of the inflection point of the curve (n =6) as the optimal number of factors based on the rubble plot of eigen value (Extended Figure 1). This is because there is no obvious benefit to the improvement in explaining the variance by adding another factor.

Third, we grouped each risk factor into a particular clustering variable based on its factor loading (Extended Figure 2). Factor loadings quantify the extent of association between variables and factors, using the recommended cut-off values: 0.32 (poor), 0.45 (fair), 0.55 (good), 0.63 (very good), or 0.71 (excellent). Considering the clinical significance of the factors when generalized corresponding variables, we re-categorized CVD to factor 2.

Forth, to verify the goodness of fit of the model, confirmatory factor analysis (CFA) with maximum likelihood estimation was subsequently conducted^3^. Root Mean Square Error of Approximation (RMSEA) and Standardized Root Mean Square Residual (SRMR) are used to assess how well the model fits the actual observed data. Values typically range between 0 and 1, with closer to 0 indicating a better model fit and closer to 1 indicating a poorer fit. CFI (Comparative Fit Index) is used to assess how well the current model fits the baseline model. CFI values range from 0 to 1, with closer to 1 indicating a better model fit to the baseline model and closer to 0 indicating a poorer fit. Based on our categorization, the 6-factor model fit well (RMSEA = 0.042, 95%CI: 0.04-0.044; SRMR = 0.035; CFI = 0.94).

Supplementary Method 3. Consensus clustering algorithm

Consensus clustering, a prevalent technique for unsupervised clustering in high-dimensional data, has garnered considerable usage for its advantage of maintaining high cluster consensus when clustering items.

We specified a range of cluster numbers, K=2-12, and for each value of K, the consensus clustering algorithm was iteratively applied. In each iteration, a random subset comprising 80% of the data records was created without replacement, and this process was repeated 1000 times. Within each random subset, the hierarchical algorithm (based on Pearson distance) was executed, assigning each individual to one of the clusters. Upon completion of the 1000 iterations, the frequencies of pairs of individuals being clustered together were calculated, yielding a N◊N matrix of pairwise consensus values (N represents the sample size) under each K scenario. To determine the optimal cluster number (K), we check the cumulative distribution function (CDF) for each K (Extended Figure 3A) and relative change in area under the CDF curve comparing K and K – 1 (Extended Figure 3B).

To determine cluster membership, a hierarchical clustering algorithm was applied using the consensus matrix as a similarity measure. In the consensus matrix, consensus values ranging from 0 (indicating no clustering together) to 1 (indicating always clustering together) were depicted with shades from white to blue (Extended Figure 4). The consensus matrix was ordered based on the consensus clustering and presented as a dendrogram above the heatmap. The cluster memberships were indicated by colored rectangles positioned between the dendrogram and heatmap, with a corresponding legend provided above the graphic.

The determination of the optimal cluster number was referred to consensus matrix heatmap and cumulative distribution function (CDF). The CDF plot illustrated the area under the CDF curve for each K value, highlighting the point at which the CDF reached its approximate maximum. This K value indicated the number of clusters at which the consensus and cluster confidence were maximized. Additionally, the comparison of the relative change in the area under the CDF curve between K and K-1 provided further insights into the suggested optimal number of clusters.

K of 6 was determined as the optimal for the clustering model, since the negligible population-distribution effect by increasing the value of K. However, when dividing patients with AF into six clusters, one group had a population of 18 (Extended Figure S4), which was smaller than 30 (the minimum recommended number for each subgroup^4^). Therefore, we selected k=5 as the optimal cluster number.

1.	Simon N, Friedman JH, Hastie T, Tibshirani R. Regularization Paths for Cox's Proportional Hazards Model via Coordinate Descent. *Journal of Statistical Software* 2011;39:1 - 13. doi: 10.18637/jss.v039.i05

2.	Browne MW. An Overview of Analytic Rotation in Exploratory Factor Analysis. *Multivariate Behavioral Research* 2001;36:111-150. doi: 10.1207/S15327906MBR3601_05

3.	Rosseel Y. lavaan: An R Package for Structural Equation Modeling. *Journal of Statistical Software* 2012;48:1 - 36. doi: 10.18637/jss.v048.i02

4.	Dalmaijer ES, Nord CL, Astle DE. Statistical power for cluster analysis. *BMC Bioinformatics* 2022;23:205. doi: 10.1186/s12859-022-04675-1

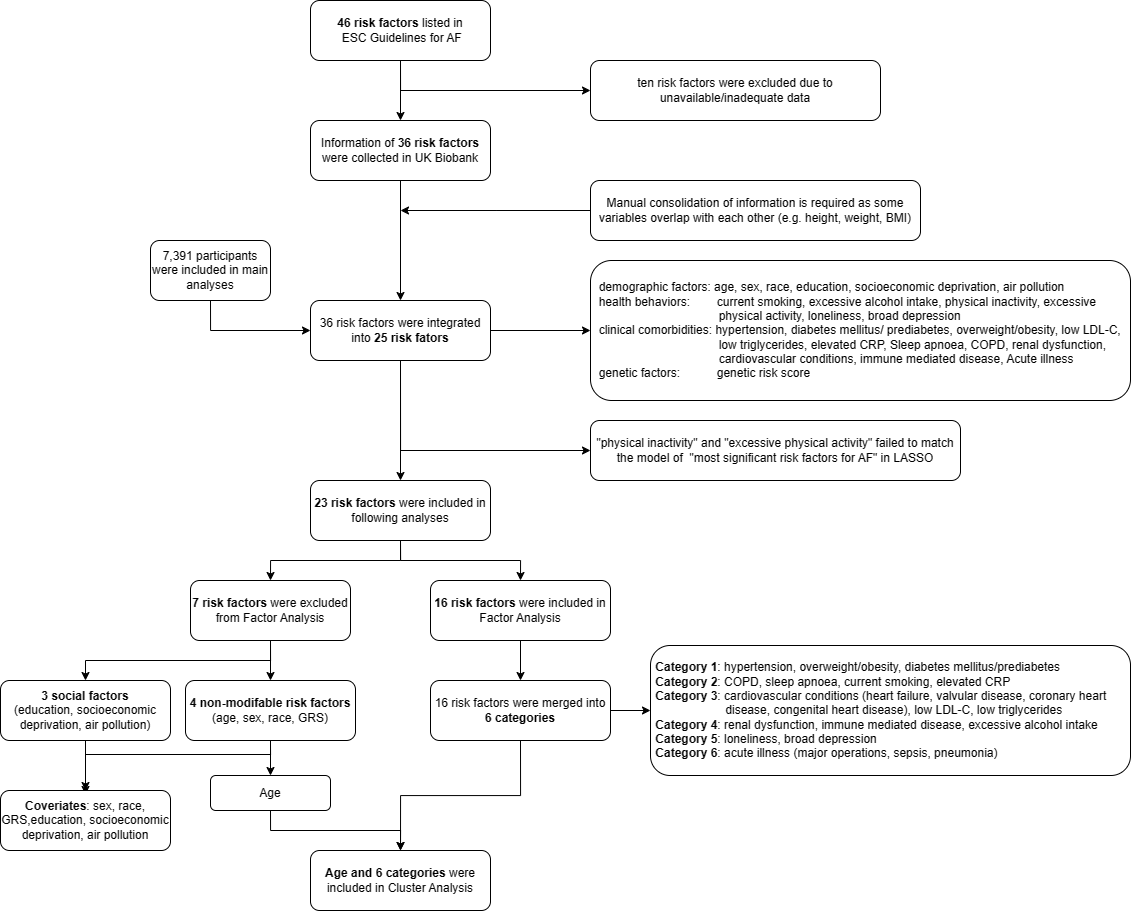

Supplementary Figure S1. Study design

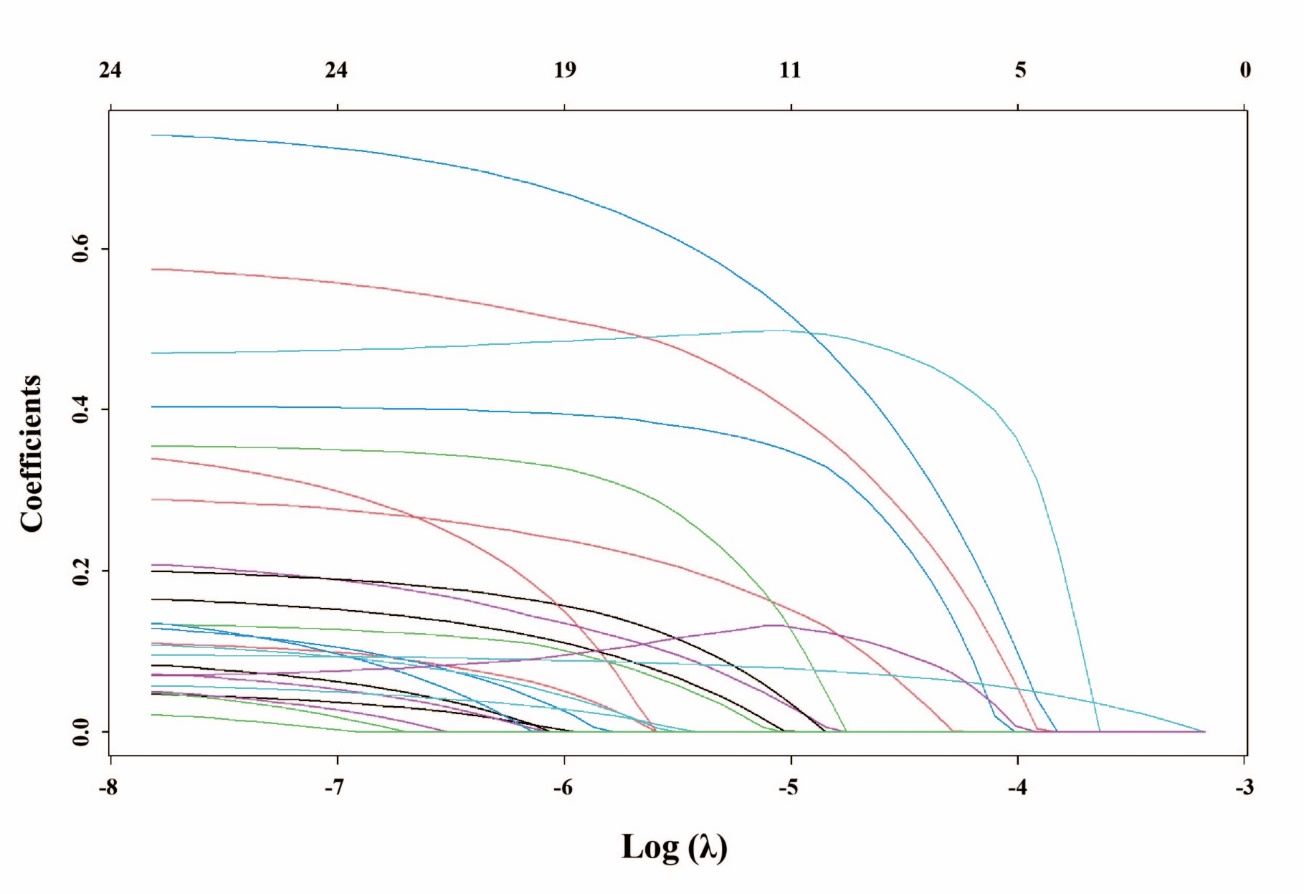
Supplementary Figure S2. Number of variables included in models for different values of lambda and the estimates of corresponding variables.

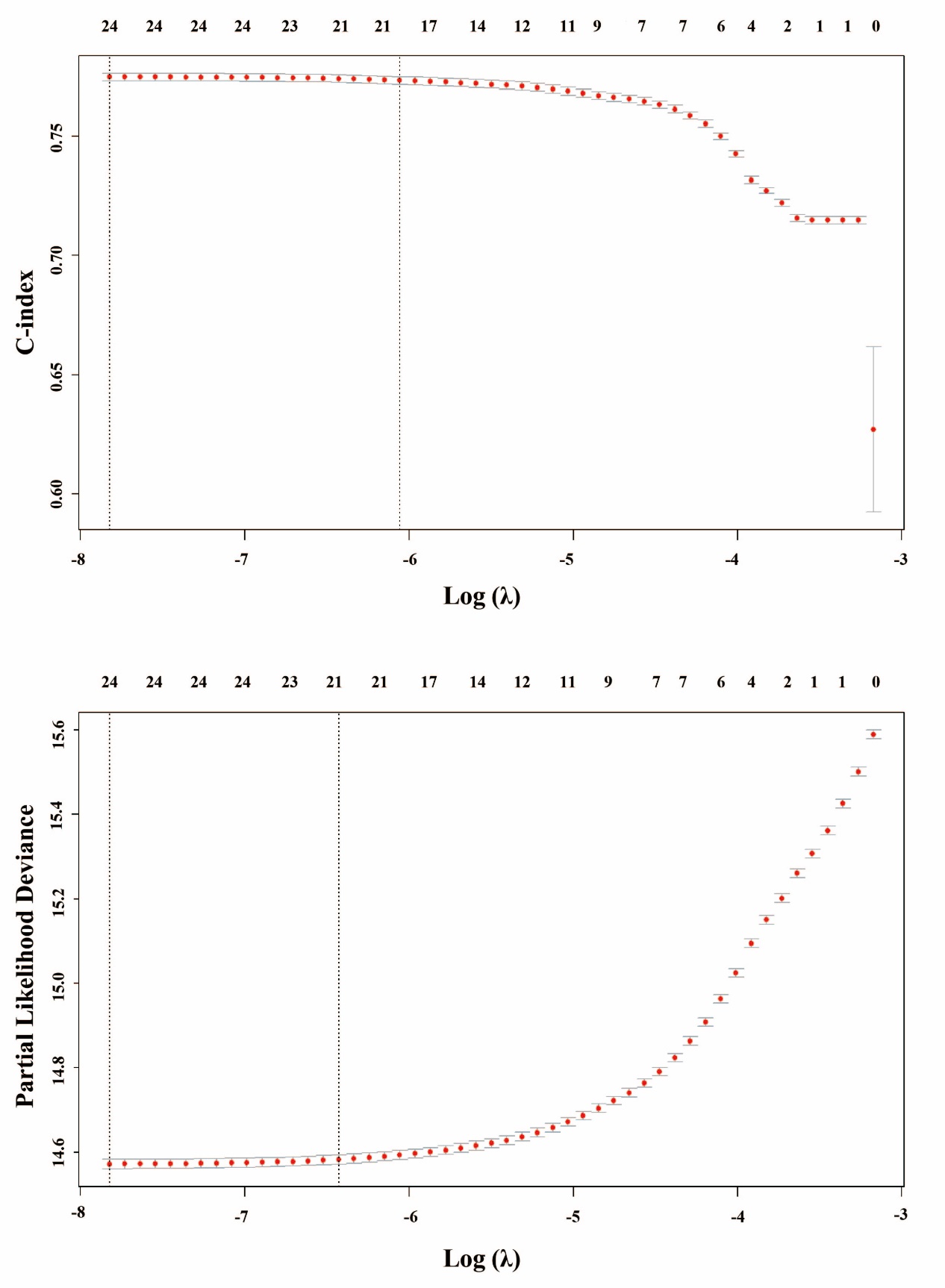

Supplementary Figure S3. The C-index and the partial likelihood deviation of the models for different values of lambda.

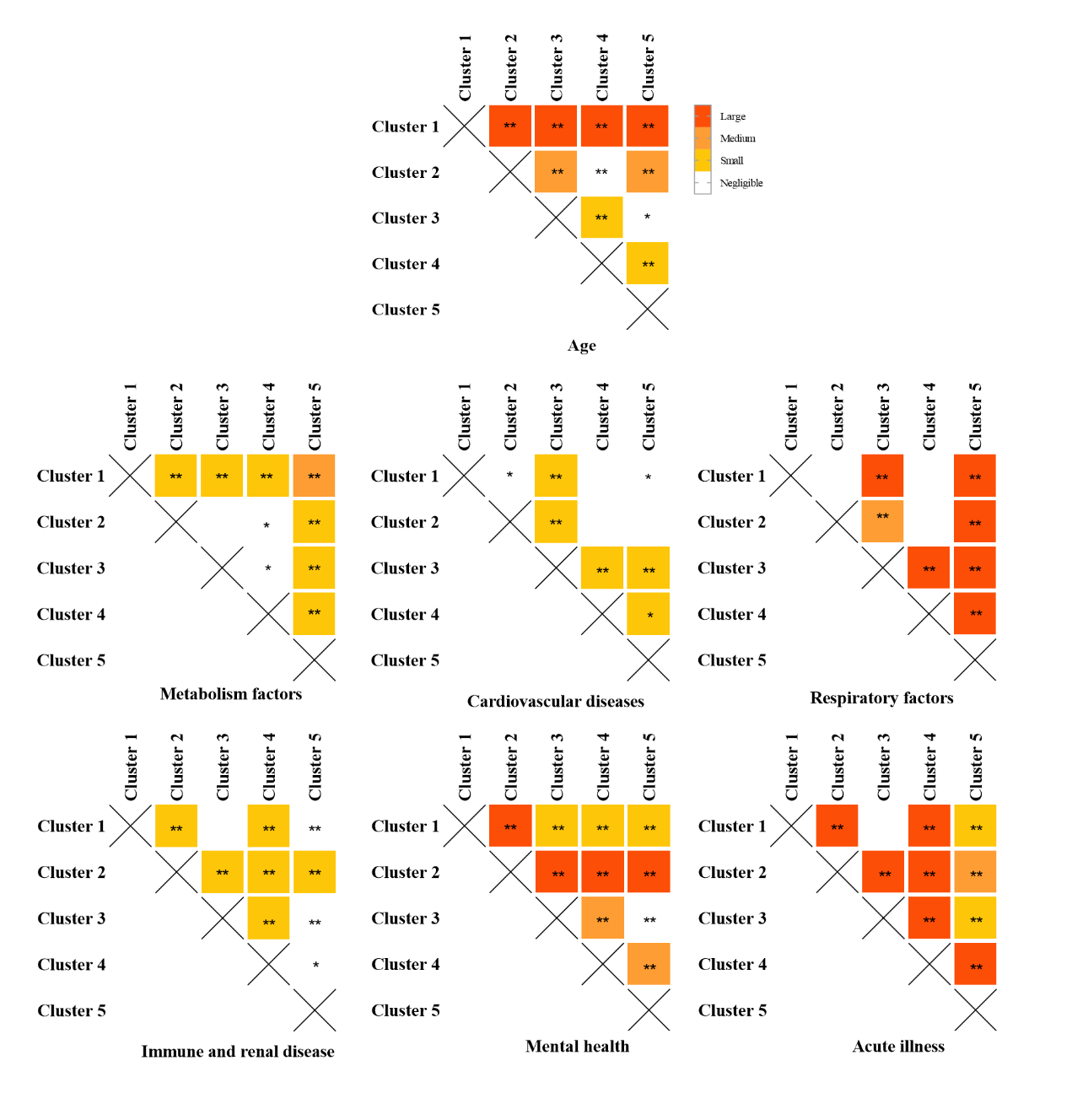

Supplementary Figure S4. Pairwise comparisons of the cluster feature variables. Bonferroni correction was applied with p < 0.005 (0.05/10) as statistical significance. The color presented the Cohen’s d values that indicated the standardized difference between the two means. Large: ≥ 0.8; medium: 0.5~0.79; small: 0.2~0.49; negligble: < 0.2.

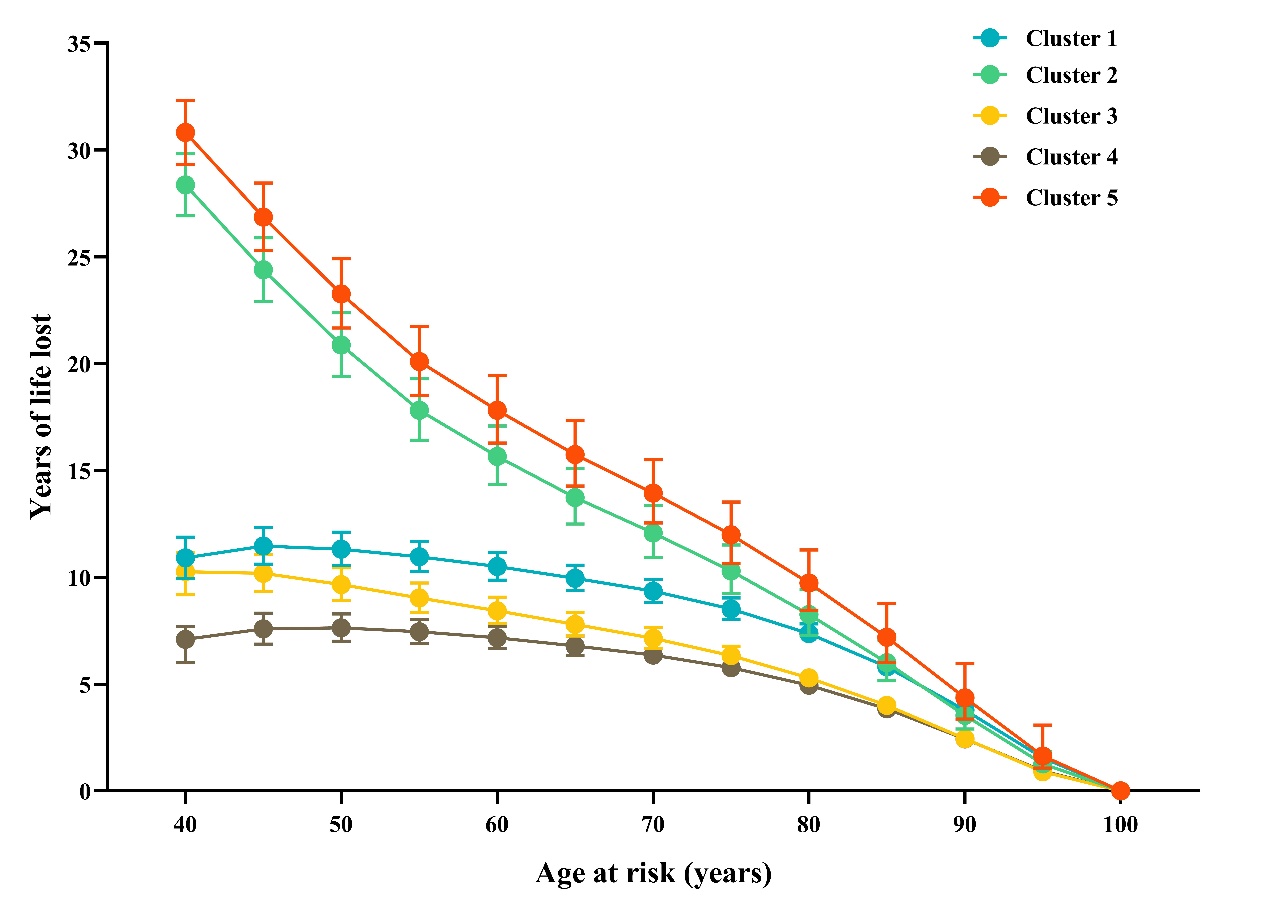

Supplementary Figure S5. Reductions of life expectency for participants in the five AF clusters comparing with non-AF participants.

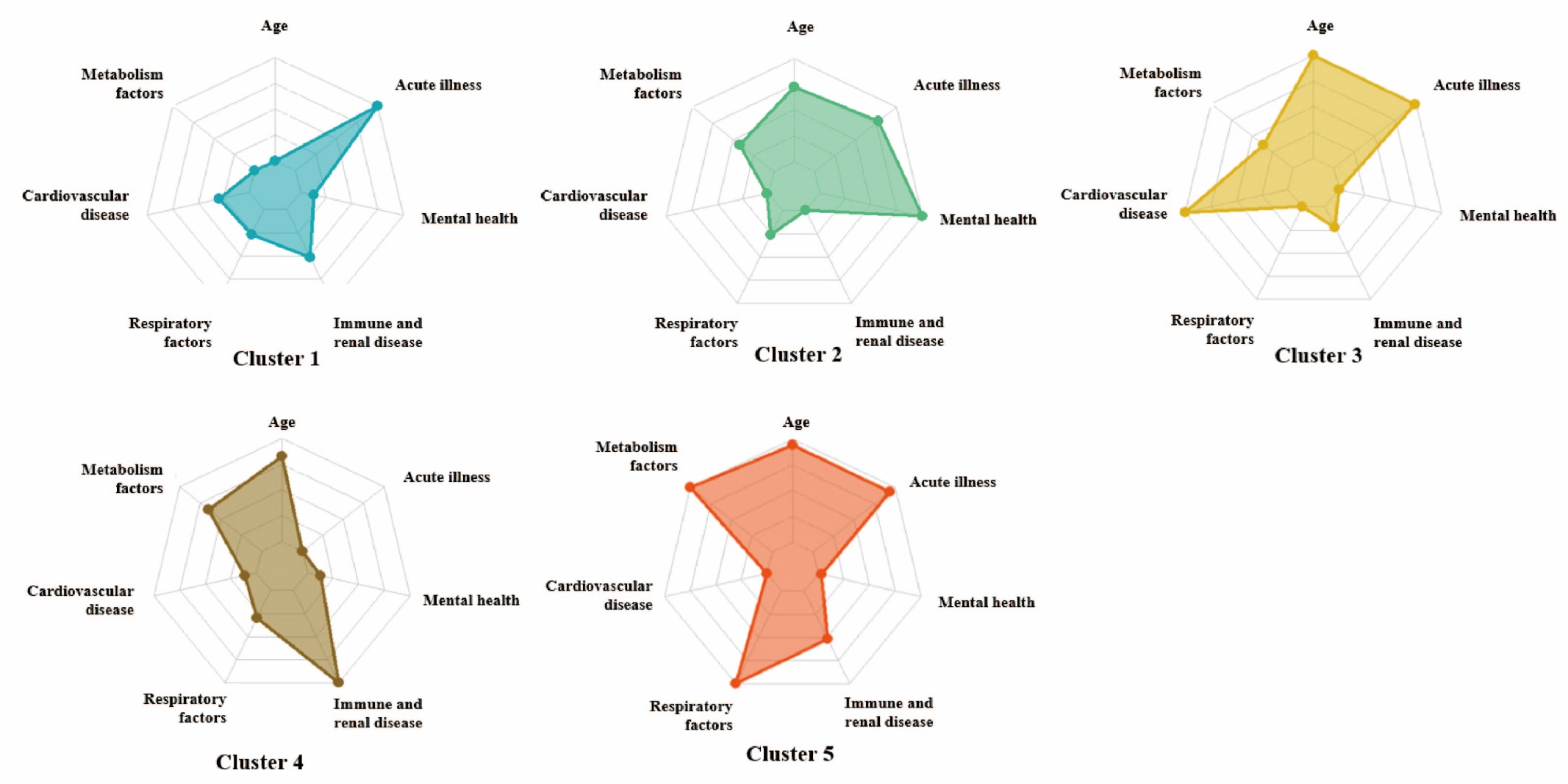

Supplementary Figure S6. Cluster features differed among five clusters in re-clustering analysis among the participants with AF before baseline.

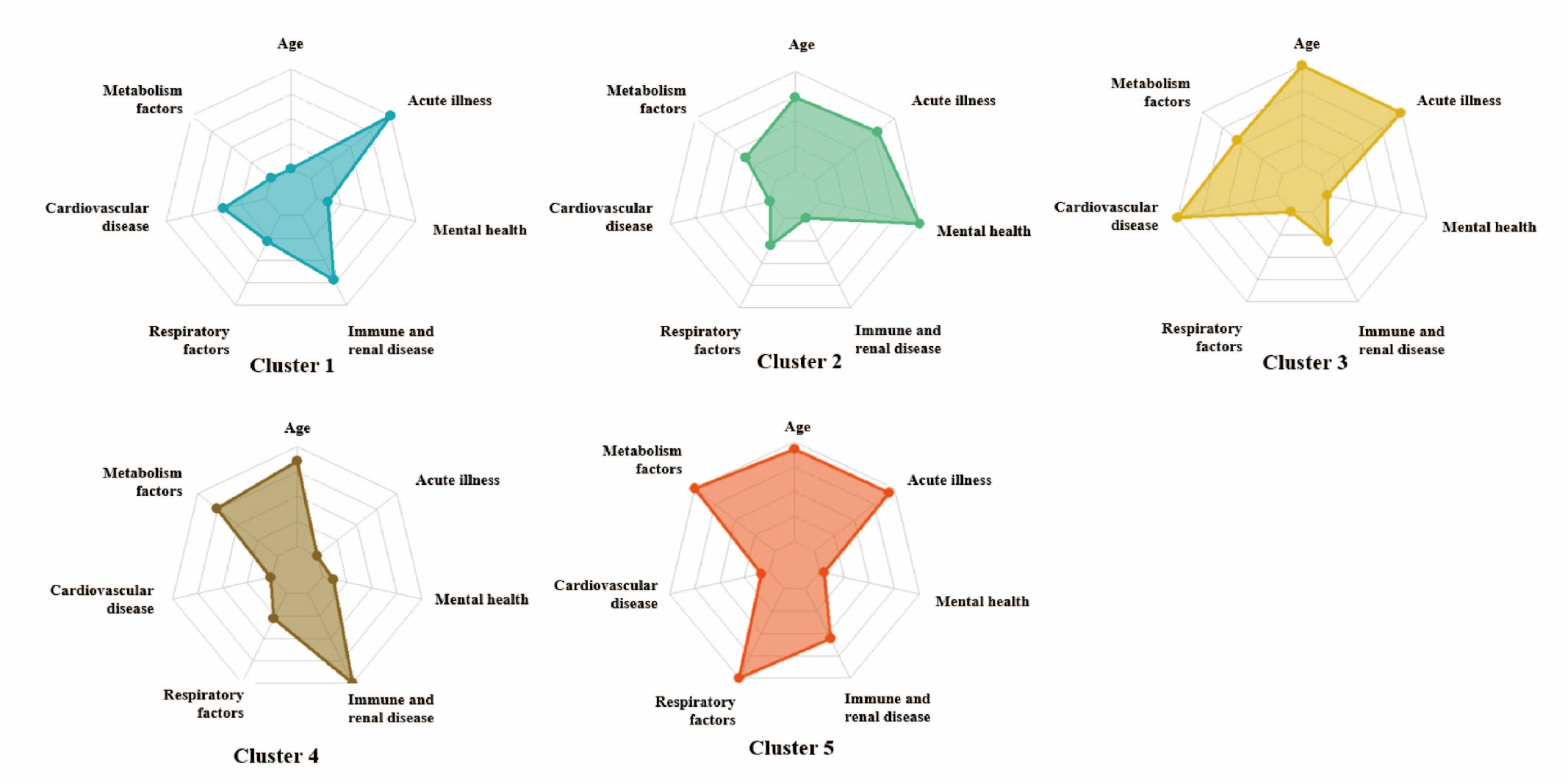

Supplementary Figure S7. Cluster features differed among five clusters under new algorithm among the participants with newly-onset AF within 2 years after baseline.

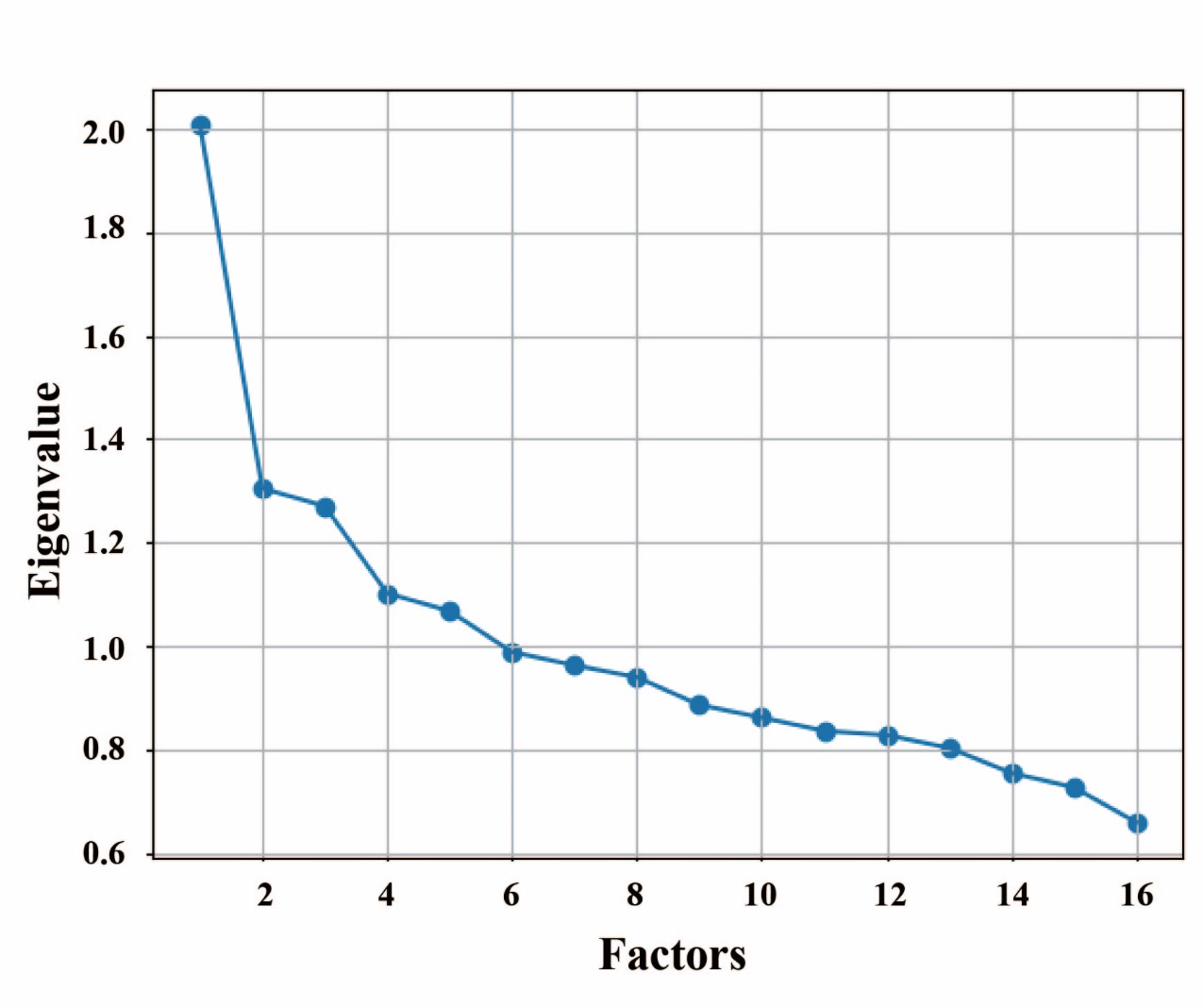

Extended Figure 1. The rubble plot of eigen value based on different numbers of factor.

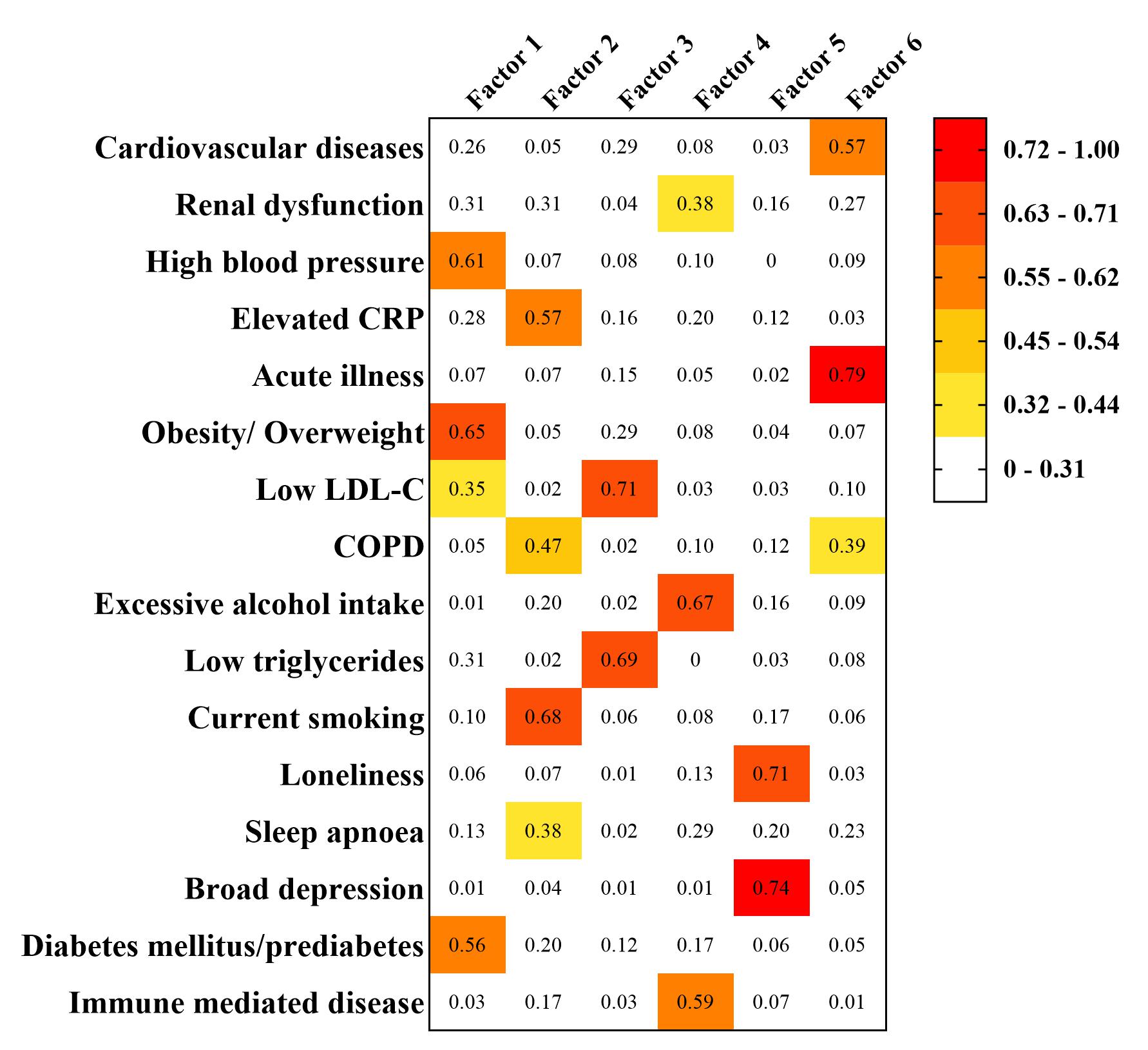

Extended Figure 2. The factor loading of 16 risk factors in 6 factors.

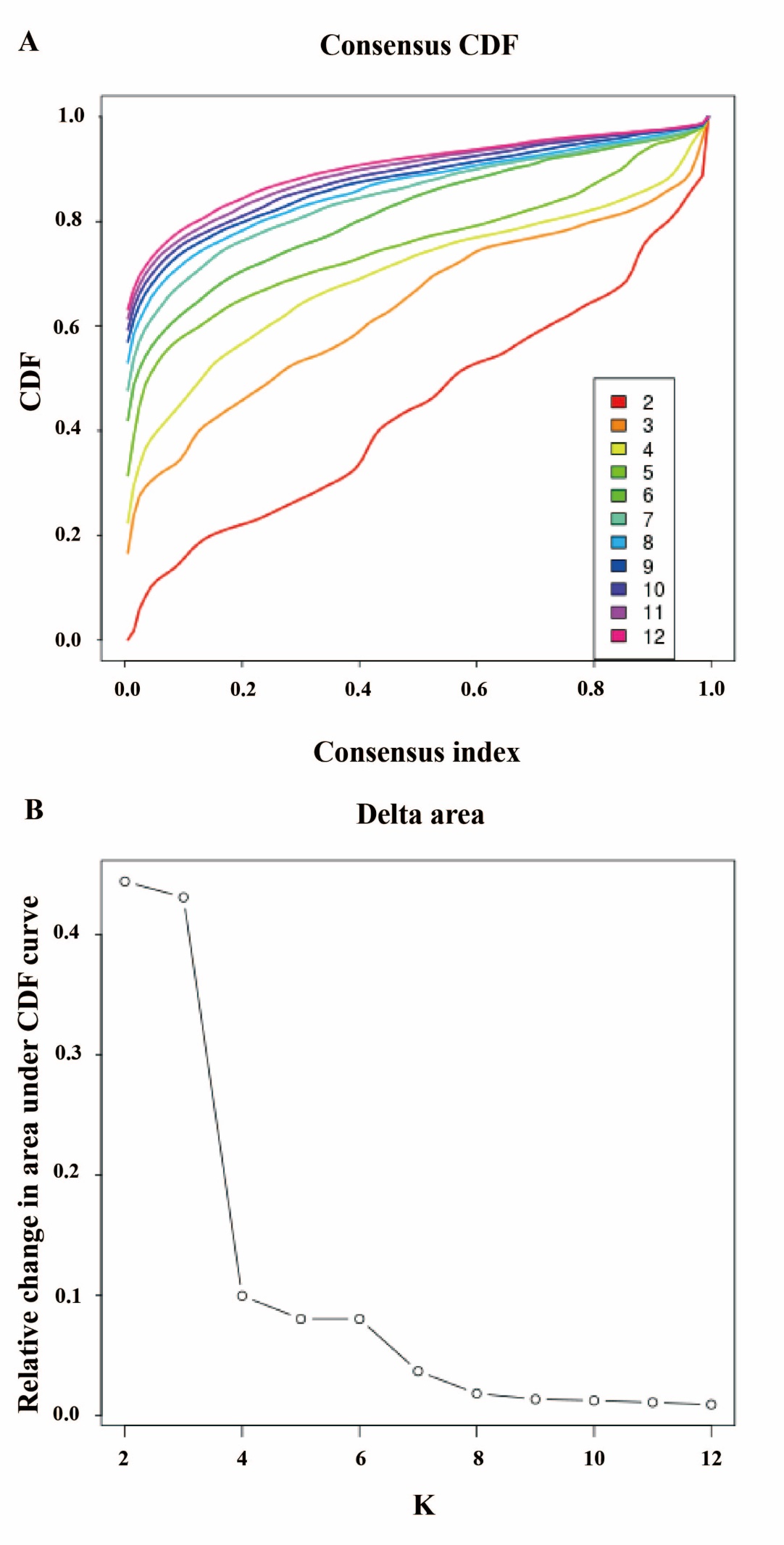

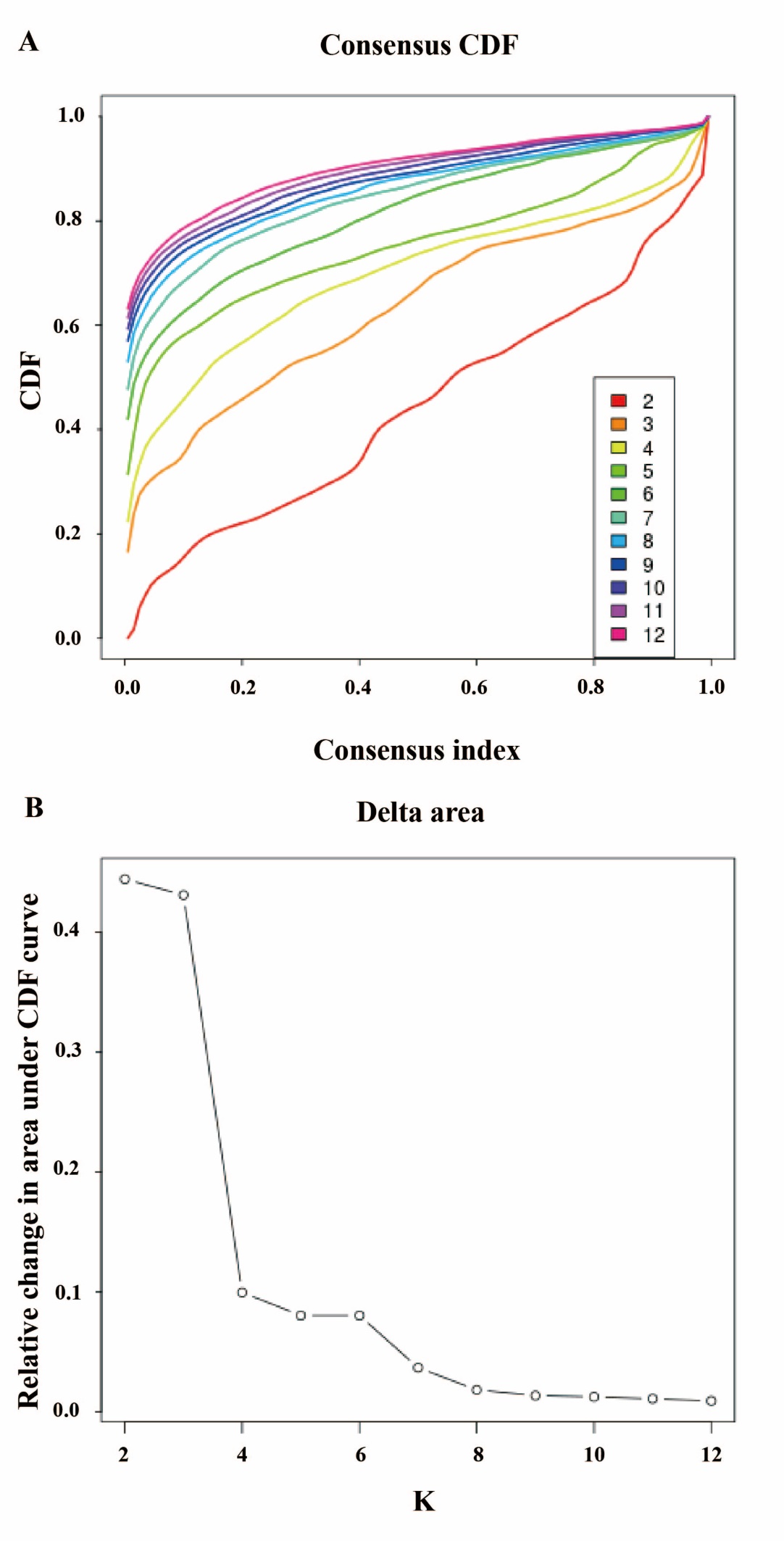

Extended Figure 3. (A) The cumulative distribution function (CDF) for each cluster number (K). (B) Relative change in area under the CDF curve comparing K and K – 1.

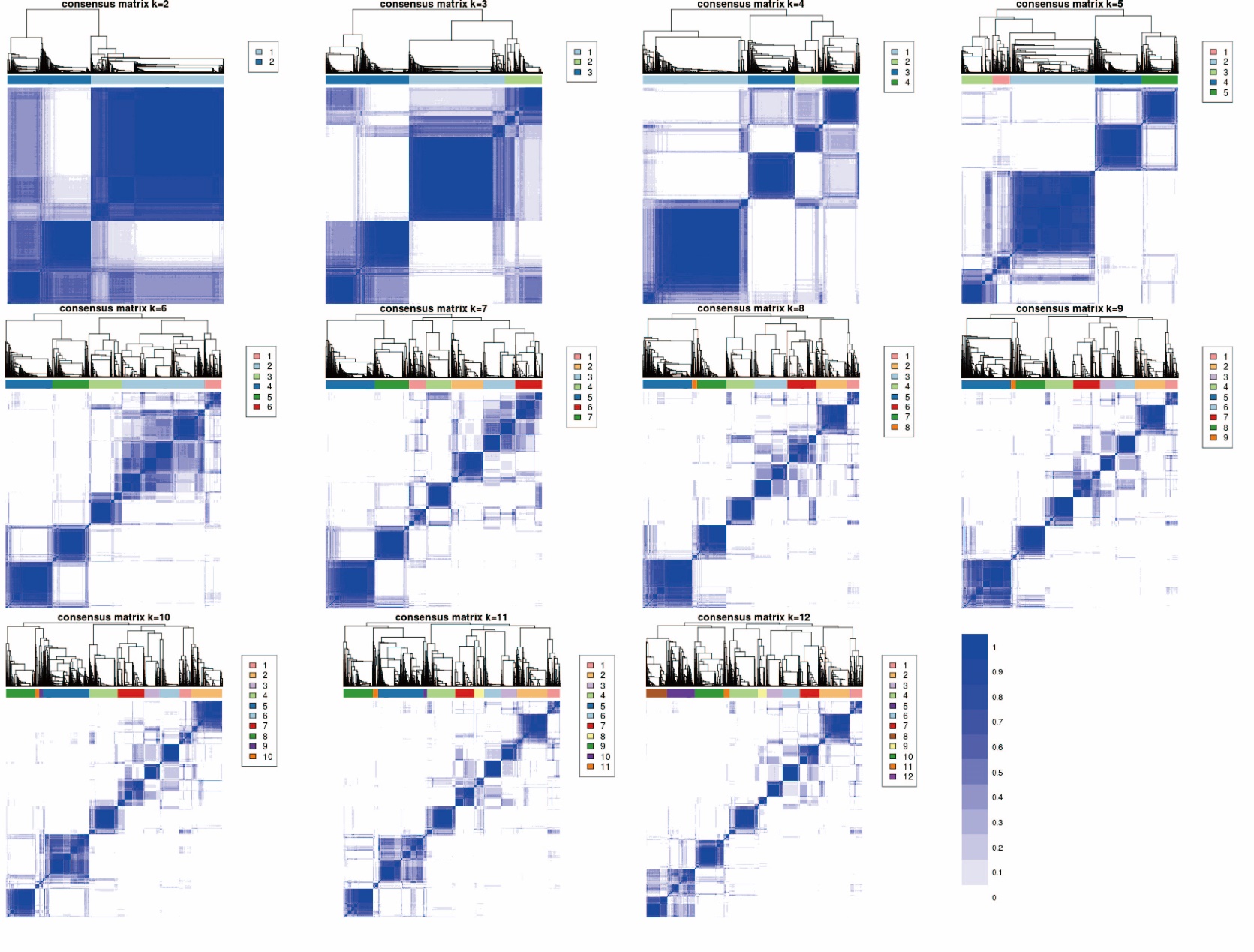

Extended Figure 4. Consensus matrix of probability of each participant categorized into the same cluster with other participants. Consensus values ranging from 0 (indicating no clustering together) to 1 (indicating always clustering together) were depicted with shades from white to blue.
